# Plasma metabolites associated with cognitive function across race/ethnicities affirming the importance of healthy nutrition

**DOI:** 10.1101/2022.03.09.22271910

**Authors:** Einat Granot-Hershkovitz, Shan He, Jan Bressler, Bing Yu, Wassim Tarraf, Casey M. Rebholz, Jianwen Cai, Queenie Chan, Tanya P. Garcia, Thomas Mosley, Bruce S. Kristal, Charles DeCarli, Myriam Fornage, Guo-Chong Chen, Qibin Qi, Robert Kaplan, Hector M. Gonzalez, Tamar Sofer

**Affiliations:** Division of Sleep and Circadian Disorders, Brigham and Women’s Hospital, Boston, MA, USA; Department of Medicine, Harvard Medical School, Boston, MA, USA; Department of Biostatistics, Harvard T.H. Chan School of Public Health, Boston, MA, USA; Human Genetics Center, School of Public Health University of Texas Health Science Center at Houston, Houston, TX, USA; Institute of Gerontology, Wayne State University, Detroit, MI, USA; Department of Epidemiology, Johns Hopkins Bloomberg School of Public Health, Baltimore, MD, USA; Department of Biostatistics, Gillings School of Global Public Health, University of North Carolina, Chapel Hill, CA, USA; Department of Epidemiology and Biostatistics, School of Public Health, Imperial College London, London, UK; Department of Neurology, School of medicine, University of Mississippi Medical Center, Jackson, MS, USA; Alzheimer’s Disease Center, Department of Neurology, University of California, Davis, Sacramento, CA, USA; Brown Foundation Institute of Molecular Medicine, McGovern Medical School, The University of Texas Health Science Center at Houston, Houston, TX, USA; Department of Epidemiology & Population Health, Albert Einstein College of Medicine, Bronx, NY, USA; Department of Nutrition and Food Hygiene, School of Public Health, Soochow University, Suzhou, China; Division of Public Health Sciences, Fred Hutchinson Cancer Research Center, Seattle WA, USA; Department of Neurosciences and Shiley-Marcos Alzheimer’s Disease Center, University of California, San Diego, La Jolla, CA, USA

**Keywords:** Metabolites, Global cognitive function, Puerto Rican, U.S. Hispanics/Latinos, Race/Ethnicities, Mediterranean diet

## Abstract

**INTRODUCTION:** We studied the replication and generalization of previously identified metabolites potentially associated with global cognitive function in multiple race/ethnicities and assessed the contribution of diet to these associations.

**METHODS:** We tested metabolite-cognitive function associations in U.S.A. Hispanic/Latino adults (n= 2,222) from the Community Health Study/ Study of Latinos (HCHS/SOL) and in European (n=1,365) and African (n=478) Americans from the Atherosclerosis Risk In Communities (ARIC) Study. We applied Mendelian Randomization (MR) analyses to assess causal associations between the metabolites and cognitive function and between Mediterranean diet and cognitive function.

**RESULTS:** Six metabolites were consistently associated with lower global cognitive function across all studies. Of these, four were sugar related (e.g., ribitol). MR analyses provided weak evidence for a potential causal effect of ribitol on cognitive function and bi-directional effects of cognitive performance on diet.

**DISCUSSION:** Several diet-related metabolites were associated with global cognitive function across studies with different race/ethnicities.

## 1. Background

Poor global cognitive function in older adults is a risk marker for morbidity and mortality[1]. In the last decade, metabolome profiling has emerged as a new approach for biomarker discovery, with metabolites used to evaluate the pathophysiology of health and disease[2]. Several studies have shown blood metabolite associations with cognitive function and dementia[3,4]. A recent study identified 13 metabolites potentially associated with either higher (3 metabolites) or lower (10 metabolites) global cognitive function in the Boston Puerto Rican Health Study (BPRHS), a cohort of 736 Puerto Rican older adults[5]. Many of the metabolites identified in the BPRHS implicate glycemic control and insulin resistance as key factors linked to cognition[5]. They also constructed a metabolite risk score, combining several metabolites as a single biomarker that could potentially discriminate good from poor extremes of cognition. Replication of these results in independent datasets is needed in order to validate the associations and to estimate more accurate effect size estimates, that do not suffer from the Winner’s curse[6]. It is also important to study whether these associations are indeed specific to Puerto Ricans, or whether they are more generally applicable to other populations of varied race/ethnicities. Blood metabolites are influenced by genetics, health conditions, lifestyle, and environmental factors such as diet[7], all of which vary, potentially resulting in group-specific associations.

Diet is an important source for many of the metabolites identified by BPRHS, specifically fruits, and vegetables, which are major components of the Mediterranean diet[8]. Adherence to the Mediterranean diet has previously been associated with cognitive benefits[9]. The BPRHS findings suggest that a healthy diet, rich in fruits and vegetables, may have a potential causal effect on cognition via a mediating role of metabolites. The opposite may also hold; cognitive function determines the diet which affects the blood metabolites.

Here, we studied whether the associations reported by the BPRHS replicate and generalize to other race/ethnicities. We also sought evidence for causal effects of the replicated metabolites on global cognitive function, as well as evidence for causal effects of diet, a major determinant of the metabolites, on global cognitive function. Our investigation followed the steps in Figure 1. First, we used data from the Hispanic Community Health Study/ Study of Latinos (HCHS/SOL), a population-based longitudinal cohort study of Hispanics/Latino adults in the U.S. who self-identified with six Hispanic/Latino background groups: Cuban, Dominican, Puerto Rican, Mexican, Central American, and South American[10]. We tested whether single metabolite associations reported in BPRHS replicate in HCHS/SOL Puerto Ricans and generalize more broadly to Hispanics/Latinos (Step A). We also assessed whether the metabolite risk score (MRS) developed in BPRHS is associated with extremes of global cognition (Step B). Next, we further assessed whether metabolite associations with global cognitive function generalize to European and African Americans using the Atherosclerosis Risk in Communities (ARIC) study (Step C). We then applied two-sample Mendelian randomization (MR) to test whether the associated metabolites are causally associated with global cognitive function (Step D). We then studied the role of diet in the observed associations between metabolites and global cognitive function. We first tested the association between the Mediterranean diet and single metabolites (Step E) and then evaluated cause-and-effect relationships between the Mediterranean diet and cognitive performance using bidirectional MR (Step F).

**Figure 1:**
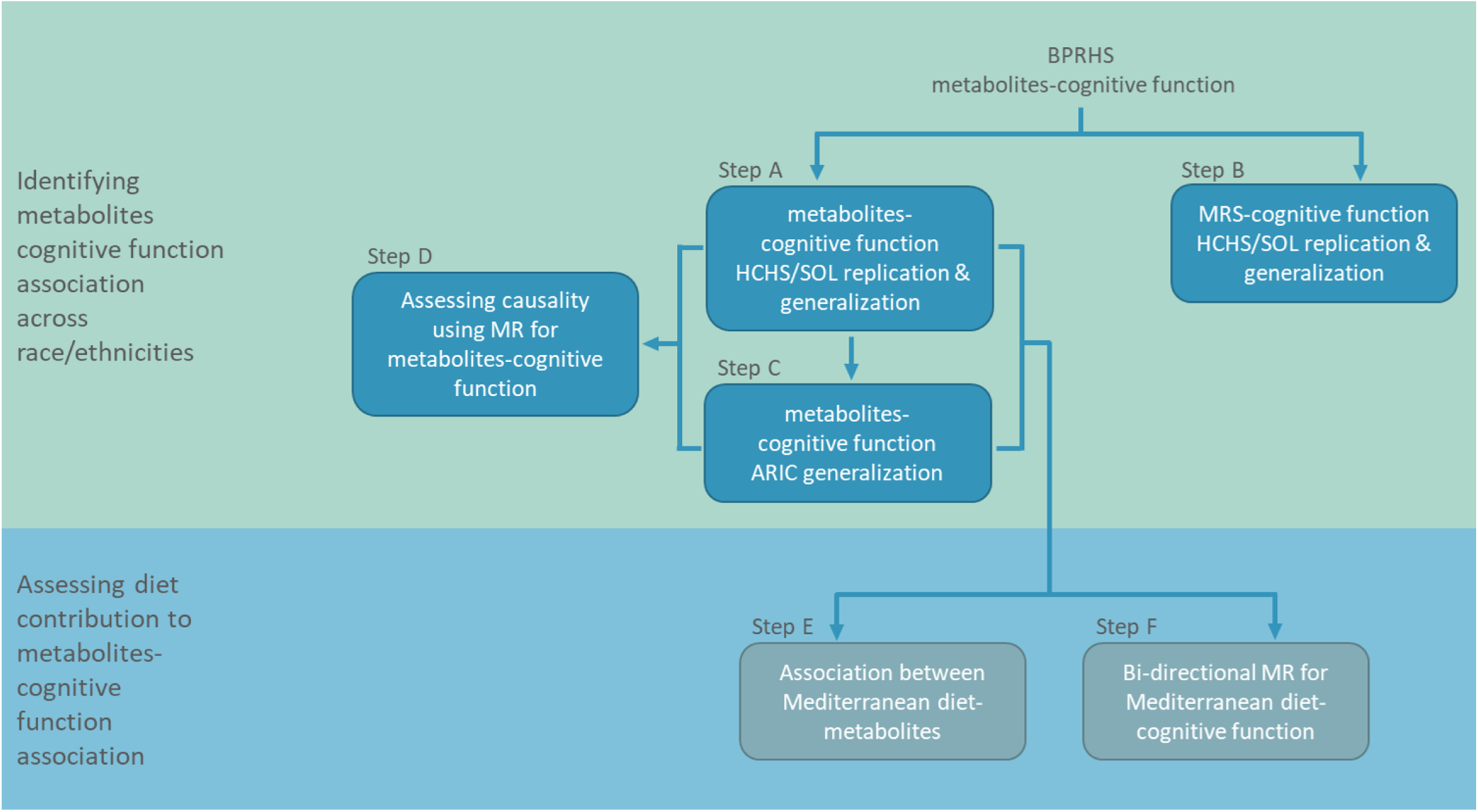
Metabolite - global cognitive function analyses flowchart. Step A: Replication and generalization analysis in HCHS/SOL of previously reported single metabolites associated with global cognitive function in BPRHS (Step A). Step B: Replication and generalization testing in HCHS/SOL of association of the metabolite risk score (MRS) constructed in BPRHS with extremes of global cognition. Step C: Generalization of single metabolite associations with global cognitive function in the Atherosclerosis Risk In Communities (ARIC) dataset. Step D: Two-sample Mendelian randomization (MR) analysis to evaluate causal relationships between the single metabolites and global cognitive function. Step E: Associations between the Mediterranean diet and single metabolites. Step F: Bidirectional MR to understand the cause-and-effect relationships between the single-food intake scores and global cognitive function.

## 2. Methods

### 2.1 HCHS/SOL Study population

The HCHS/SOL is a population-based longitudinal multisite cohort study of Hispanic/Latino adults in the

U.S. that enrolled participants from primarily six self-identified backgrounds: Cuban, Central American, Dominican, Mexican, Puerto Rican, and South American[10,11]. A total of 16,415 adults, 18–74-year-old, were enrolled at baseline visit 1 at four field centers (Bronx, NY, Chicago, IL, Miami, FL, and San Diego, CA) (2008-2011). A detailed description of the sampling design, including the generation and use of survey weights for the HCHS/SOL, was previously published[10,11]. Anthropometry, biospecimens, and health information about risk/protective factors were collected. Cognitive function was assessed in 9,714 individuals aged ≥45 years via a battery of 4 tests: Six-Item Screener (SIS; mental status)[12]; Brief-Spanish English Verbal Learning Test (B-SEVLT; verbal episodic learning and memory)[13]; Word Fluency (WF; verbal functioning)[14]; and Digit Symbol Substitution test (DSS; processing speed, executive function)[15]. Metabolites were measured in serum, after fasting, on a random subset of 3,978 participants from visit 1, of which 2,366 also had cognitive data. We included individuals with cognitive data, with <25% missing measured metabolites, with genetic consent, with *APOE* genotype and no missing covariates. Overall, our HCHS/SOL analytic sample included 2,222 participants, of which 426 were of Puerto-Rican background.

The details of the IRB/oversight body that provided approval or exemption for the research described are given below:

#### Mass General Brigham IRB

All necessary patient/participant consent has been obtained and the appropriate institutional forms have been archived.

### 2.2 Neurocognitive outcome

We mimicked the analysis performed in BPRHS[5] and calculated a global cognitive function score as the mean of the z-scores for each of the available cognitive tests: SIS (rank normalized prior to z-score computation to achieve more symmetric distribution compared to its raw values), WF, DSS, and two B-SEVLT scores (recall and delayed recall). We determined good and poor extremes of global cognition by including individuals with ≤1SD below the mean and individuals with ≥1SD above the mean, totaling in n=172 Puerto Ricans (109 high and 63 low global cognition scores) and n=724 in the total HCHS/SOL analytical sample (361 high and 363 low global cognition scores).

### 2.3 Metabolomics measurement and processing

Metabolites were measured in blood serum drawn after ≥8 hours of overnight fasting. Profiling was done using untargeted liquid chromatography-mass spectrometry (LC-MS) using the discovery HD4 platform in 2017 at Metabolon Inc. (Durham, NC)[16]. Out of 13 previously identified single metabolite associations with global cognitive function in the BPRHS, 11 metabolites were available in the HCHS/SOL (Supplementary Table 1). Out of 51 metabolites included in model 1 MRS associated with global cognitive function in the BPRHS, 36 metabolites were available in the HCHS/SOL (Supplementary Table 2). MRS for the HCHS/SOL participants was calculated based on these metabolites and their concordant estimated effects. Metabolites were treated as continuous measures and rank-based inverse normalized across the sample. All identified metabolites had <26% missing values. Missing values were imputed with half the minimum observed value (that is not zero) in the sample.

### 2.4 Genotyping

*APOE* genotyping was performed using commercial TaqMan assays previously described[17]. For individuals with missing *APOE* genotypes, we determined *APOE* genotypes based on phased whole-genome sequencing (WGS) data from TOPMed Freeze 8. Other genetic data were used based on genotyping (rather than WGS) using an Illumina custom array, previously reported. Principal Components (PCs) were previously computed using PC-Relate[18], and the kinship matrix was computed using the genetic data. ‘Genetic analysis groups’ were constructed based on a combination of self-identified Hispanic/Latino background and genetic similarity, and are classified as Central American, Cuban, Dominican, Mexican, Puerto Rican, and South American[19]. Genome-wide imputation was conducted using the TOPMed Freeze 5b as a reference panel[20].

### 2.5 Statistical Analyses

We characterized our HCHS/SOL analytic sample stratified by Hispanic/Latino background.

#### 2.5.1 Replication and generalization of metabolite and MRS associations in HCHS/SOL

We tested the cross-sectional association between each of the 11 metabolites and MRS with global cognitive function in the HCHS/SOL Puerto-Ricans and in the total HCHS/SOL analytical sample (Figure 1, Step A). We used generalized linear mixed models with the ‘GENESIS’ R package with correlations modeled via kinship, household, and census block unit sharing matrices. We used this approach to account for the correlation structure between individuals in the sample and included genetic relatedness because some metabolites are highly heritable. Similar to the BPRHS, we adjusted for age, sex, study center, education (<12,12,12< years), BMI (normal, overweight, obese), smoking (never, past, current), *APOE*-ε4 carrier status (dominant mode), first five PCs, and Mediterranean dietary score. The Mediterranean dietary score is derived from the mean intakes of food groups/nutrients from two 24-hour dietary recalls, ranging between 0 (lowest adherence to the diet) and 9 (highest adherence to the diet) [21]. The food groups/nutrients include nine scores: legume, fruit, vegetable, wholegrain, nuts, red meat, fish, monounsaturated fatty acids (MUFA)/saturated fatty acids (SFA) ratio, and alcohol. When estimating associations in the total HCHS/SOL analytical sample, we also adjusted for self-identified Hispanic/Latino background groups. We combined previous association results for single-metabolite association with global cognitive function in BPRHS with the results in the HCHS/SOL Puerto-Ricans and total HCHS/SOL analytic sample in an inverse-variance weighted, fixed-effect meta-analysis, using one-sided p-value for HCHS/SOL results. We mimicked the analysis performed in BPRHS and tested whether the MRS could discriminate good from poor extremes of global cognition by using the generalized linear mixed models with a binomial link function adjusted for the above-listed covariates (Figure 1, Step B). An association was significant if its one-sided p-value was < 0.05.

#### 2.5.2 Sensitivity analyses

We mimicked the BPRHS sensitivity analyses and restricted the analyses to participants without diabetes (based on American Diabetes Association definition) (Puerto Rican n=295, total n=1,631) or participants with good adherence to the Mediterranean diet (diet-score 4 or above) (Puerto Rican n=143, total n=1,101) or participants aged 60 or more years (Puerto Rican n=139, total n=603) or to non-*APOE*-ε4 carriers (Puerto Rican n=317, total n=1,724).

#### 2.5.3 Generalization analysis of metabolite associations in ARIC study and meta-analysis

We further evaluated the generalizability of the discovered single metabolite associations in the ARIC study (Figure 1, Step C) which is comprised of other major U.S. race/ethnic groups, European and African Americans. ARIC is a longitudinal cohort study with cognitive measures and metabolite profiling based on a similar Metabolon platform[22,23]. A global cognitive function score was derived from the mean of the z-scores for each of the following cognitive tests administered at visit 2 (1990-1992): Delayed Word Recall Test (DWRT), Word Fluency Test (WFT), and Digit Symbol Substitution Test (DSST)[24]. Adherence to a Mediterranean diet was evaluated by calculating the alternate Mediterranean diet (aMed) score using data obtained by administering a semiquantitative food frequency questionnaire to assess dietary intake at visit 1 (1987-1989)[25,26]. Further details are provided in the Supplementary Materials and Table 1. Out of 13 previously identified single metabolite associations with global cognitive function in the BPRHS, 10 metabolites were available in ARIC. We meta-analyzed the results from BPRHS, HCHS/SOL Hispanics/Latinos, ARIC European, and African Americans in an inverse-variance, fixed-effect meta-analysis.

**Table 1:** Demographics, health, and lifestyle characteristics of the HCHS/SOL and ARIC analytic study samples, stratified by background or ancestry.

|  | HCHS/SOL |  |  |  |  |  |  |  | ARIC |  |
| --- | --- | --- | --- | --- | --- | --- | --- | --- | --- | --- |
|  | Dominican | Central American | Cuban | Mexican | Puerto-Rican | South American | More than one heritage/Other | Total | European American | African American |
| N (%) | 217 (9.8) | 213 (9.6) | 433 (19.5) | 749 (33.7) | 426 (19.2) | 136 (6.1) | 48 (2.2) | 2,222 | 1,365 | 478 |
| SEX = M (%) | 84 (38.7) | 80 (37.6) | 209 (48.3) | 286 (38.2) | 172 (40.4) | 56 (41.2) | 25 (52.1) | 912 (41.0) | 620 (45.4) | 164 (34.3) |
| AGE (years) (mean (SD)) | 53.88 (7.44) | 55.01 (7.20) | 55.43 (7.43) | 54.26 (7.29) | 56.07 (7.49) | 54.67 (7.36) | 55.04 (8.04) | 54.91 (7.41) | 57.47 (5.74) | 56.29 (5.41) |
| Education years (%) |  |  |  |  |  |  |  |  |  |  |
| <12 | 97 (44.7) | 92 (43.2) | 111 (25.6) | 357 (47.7) | 180 (42.3) | 29 (21.3) | 20 (41.7) | 886 (39.9) | 223 (16.3) | 195 (40.8) |
| 12 | 41 (18.9) | 40 (18.8) | 109 (25.2) | 154 (20.6) | 101 (23.7) | 31 (22.8) | 9 (18.8) | 485 (21.8) | 454 (33.3) | 95 (19.9) |
| >12 | 79 (36.4) | 81 (38.0) | 213 (49.2) | 238 (31.8) | 145 (34.0) | 76 (55.9) | 19 (39.6) | 851 (38.3) | 688 (50.4) | 188 (39.3) |
| BMI (kg/m2) (%) |  |  |  |  |  |  |  |  |  |  |
| Normal weight (< 25.0) | 30 (13.8) | 28 (13.1) | 93 (21.5) | 111 (14.8) | 69 (16.2) | 31 (22.8) | 6 (12.5) | 368 (16.6) | 452 (33.1) | 86 (18.0) |
| Overweight (25.0 – 30.0) | 87 (40.1) | 89 (41.8) | 178 (41.1) | 296 (39.5) | 148 (34.7) | 56 (41.2) | 12 (25.0) | 866 (39.0) | 568 (41.6) | 183 (38.3) |
| Obese (> 30) | 100 (46.1) | 96 (45.1) | 162 (37.4) | 342 (45.7) | 209 (49.1) | 49 (36.0) | 30 (62.5) | 988 (44.5) | 345 (25.3) | 209 (43.7) |
| Smoking (%) |  |  |  |  |  |  |  |  |  |  |
| Never | 145 (66.8) | 127 (59.6) | 176 (40.6) | 438 (58.5) | 206 (48.4) | 75 (55.1) | 28 (58.3) | 1,195 (53.8) | 300 (22.0) | 131 (27.4) |
| Former | 46 (21.2) | 53 (24.9) | 114 (26.3) | 206 (27.5) | 104 (24.4) | 37 (27.2) | 13 (27.1) | 573 (25.8) | 555 (40.7) | 142 (29.7) |
| Current | 26 (12.0) | 33 (15.5) | 143 (33.0) | 105 (14.0) | 116 (27.2) | 24 (17.6) | 7 (14.6) | 454 (20.4) | 510 (37.4) | 205 (42.9) |
| Mediterranean diet score (mean (SD))* | 4.45 (1.54) | 4.67 (1.56) | 4.14 (1.44) | 5.05 (1.53) | 3.85 (1.55) | 4.66 (1.55) | 4.58 (1.60) | 4.51 (1.59) | 4.25 (1.8) | 4.29 (1.7) |
| APOE -ε4 carriers (%) | 75 (34.6) | 50 (23.5) | 96 (22.2) | 142 (19.0) | 109 (25.6) | 17 (12.5) | 9 (18.8) | 498 (22.4) | 406 (29.7) | 196 (41.0) |
| Type II Diabetes (%) | 53 (24.4) | 53 (24.9) | 108 (24.9) | 215 (28.7) | 131 (30.8) | 17 (12.5) | 14 (29.2) | 591 (26.6) | 147 (10.8) | 134 (28.2) |
| Global cognitive score (mean (SD)) | -0.19 (0.70) | -0.04 (0.67) | -0.01 (0.70) | 0.17 (0.71) | -0.13 (0.74) | 0.32 (0.66) | 0.04 (0.74) | 0.03 (0.72) | 0.18 (0.7) | -0.58 (0.8) |

**Abbreviations:** *HCHS/SOL*, Hispanic Community Health Study/Study of Latinos; *ARIC*, Atherosclerosis Risk In Communities study.
\*For ARIC we used an alternate Mediterranean diet (aMed) score, Fung TT et al. Am J Clin Nutr 82:163-173, 2005.

#### 2.5.4 Mendelian Randomization analysis to assess a causal effect of metabolites on global cognitive function

We performed a two-sample MR to estimate the causal effect of each of the replicated metabolites (exposures) on global cognitive function (outcome) using the ‘TwoSampleMR’ R package[27] (Figure 1, Step D). Further details are described in the supplemental data.

#### 2.5.5 Assessment of dietary contributions to the association between metabolites and cognitive function

In the Supplementary Material, we provide information regarding a secondary analysis studying the association effect of Mediterranean diet on the replicated and/or generalized metabolite associations and the potential causal effect of diet on cognitive function and vice versa using bi-directional MR.

#### 2.5.6 Data availability

HCHS/SOL genetic, phenotypic, and metabolomics data can be obtained through the study’s Data Coordinating Center using an approved data use agreement. Information is provided in https://sites.cscc.unc.edu/hchs/. HCHS/SOL genetic and phenotypic data can also be obtained from dbGaP under accession number phs000810.v1.p1. ARIC genetic, phenotypic, and metabolomics data can be obtained through the study’s Data Coordinating Center using an approved data use agreement. Information is provided in https://sites.cscc.unc.edu/aric/distribution-agreements.

## 3. Results

Table 1 characterizes the demographic, health, and lifestyle characteristics of the study population, stratified by Hispanic/Latino background groups. Overall, the HCHS/SOL analytic dataset included 2,222 individuals (912 men; 1,310 women), with a mean age of 54.9 (SD=7.4) years at visit 1. Out of these, 426 individuals were Puerto-Rican (172 men; 254 women), with a mean age of 56.1 (SD=7.5) years at visit 1.

### 3.1 Replication, generalization, and meta-analysis of metabolite and MRS association analyses in HCHS/SOL

Figure 2 and Supplementary Table 3 summarize previous association results of the 11 metabolite associations with global cognitive function in the BPRHS, together with replication results in the HCHS/SOL Puerto-Rican background group and generalization results in the total HCHS/SOL analytic sample, and their meta-analyses with BPRHS. In general, 10 out of 11 metabolite associations had a consistent direction of estimated effects between BPRHS and HCHS/SOL Puerto-Ricans, and 9 out of 11 for the total HCHS/SOL analytic sample. Three metabolite associations, beta-Cryptoxanthin, gamma-CEHC glucuronide, and Tyramine-O-sulfate, were replicated (p-value<0.05) in HCHS/SOL Puerto Ricans. Two of these metabolites, beta-Cryptoxanthin and gamma-CEHC glucuronide, and one other metabolite, 5’-Methylthioadenosine, generalized (p-value<0.05) to the total HCHS/SOL dataset. Figure 3 visualizes the Pearson correlations between the 11 available metabolites in the HCHS/SOL analytic sample.

**Figure 2:**
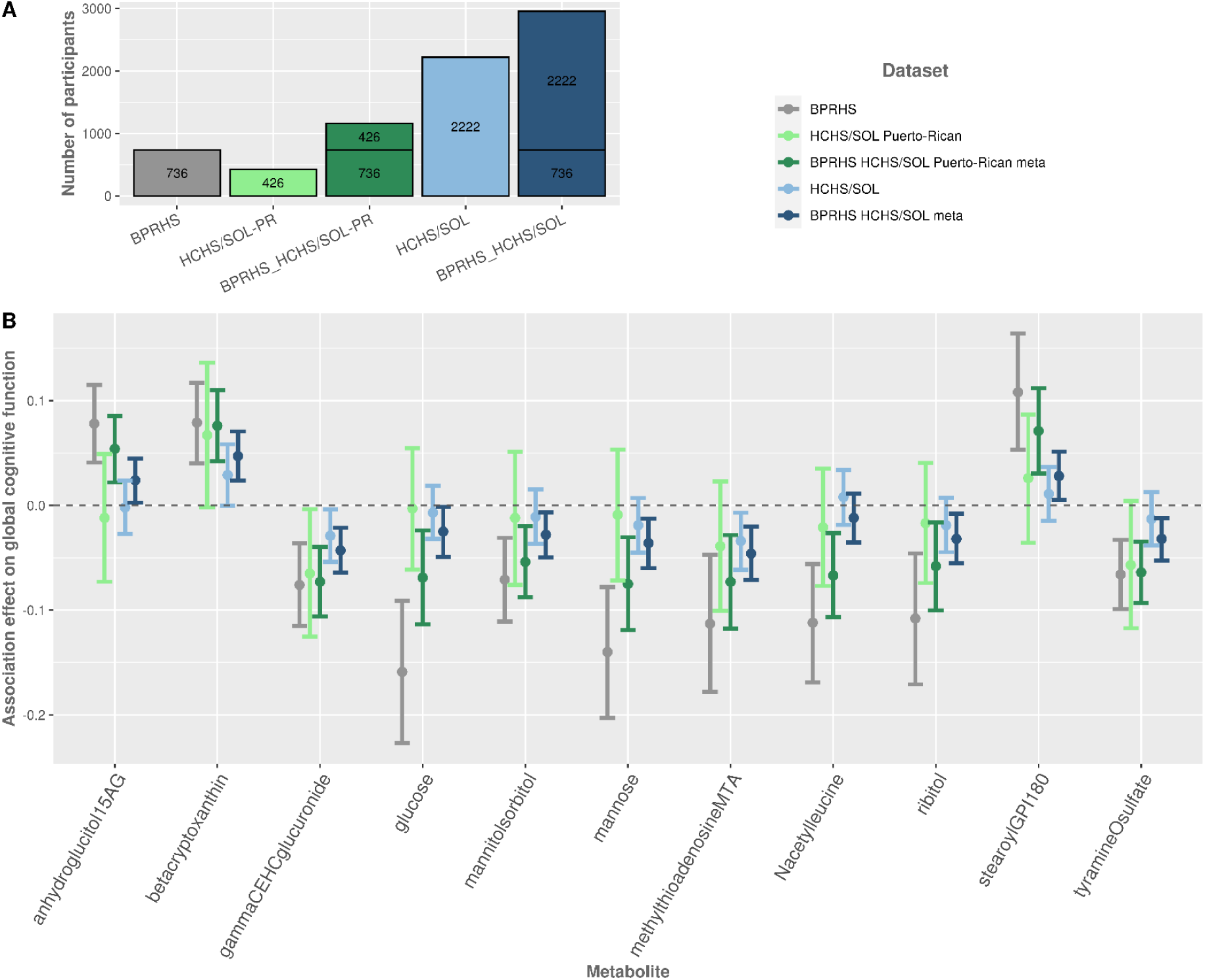
**A**. The number of participants in each study and/or combination of studies. **B**. Estimated effects and 95% confidence intervals for single metabolite associations with global cognitive function, in BPRHS (previously reported), HCHS/SOL Puerto-Ricans, and total HCHS/SOL analytical sample. Meta-analysis of the BPRHS with each of these groups is presented as well. Models are adjusted for age, sex, study center, education, BMI, smoking, *APOE*-ε4 carrier status, smoking, and Mediterranean diet score.

**Figure 3:**
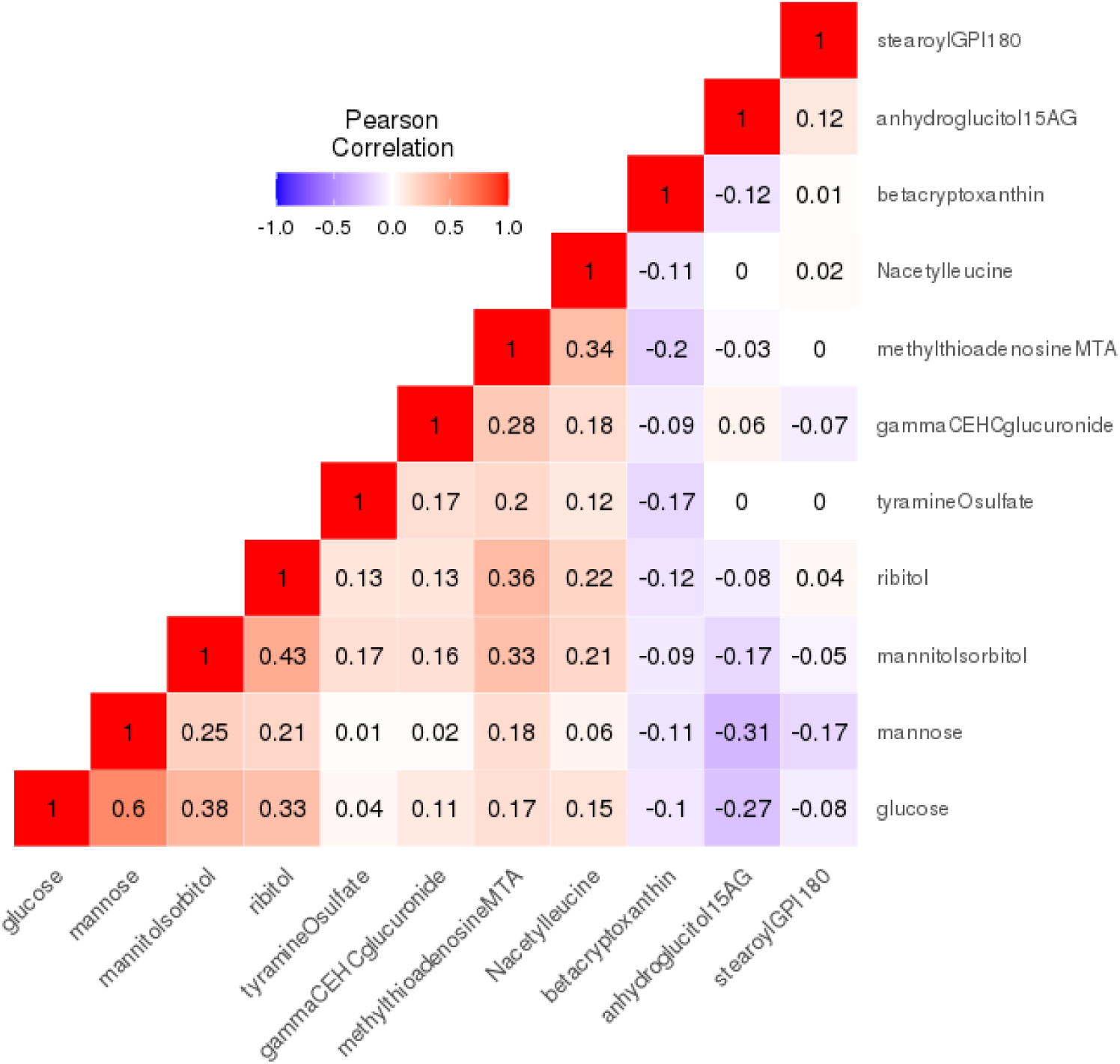
Correlations between the 11 metabolites associated with global cognitive function, previously identified in the BPRHS, and available in the HCHS/SOL. The metabolite correlations were computed within the HCHS/SOL current analytic sample (n=2,222).

The MRS, based on 36 identified metabolites out of 51 metabolites included in the BPRHS MRS, did not significantly discriminate good from poor extremes of global cognition, neither in the Puerto Rican from HCHS/SOL (OR=1.28, 95% confidence interval=[0.77,2.13]) nor in the total HCHS/SOL analytic samples (beta=1.16, 95% confidence interval=[0.92,1.46]).

### 3.2 Sensitivity analyses

In sensitivity analyses restricted to non-diabetic participants, both in the HCHS/SOL Puerto-Rican and the total HCHS/SOL analytic sample, the association of betacryptoxanthin with higher cognitive function strengthened, similar to the sensitivity results in BPRHS. It also strengthened when restricting the HCHS/SOL Puerto Ricans to non-carriers of *APOE*-ε4 or participants age 60 and above, and when restricting the total HCHS/SOL analytic sample to participants with high adherence to the Mediterranean diet or participants age 60 and above. In sensitivity analyses restricted to HCHS/SOL Puerto-Rican non-carriers of *APOE*-ε4, glucose was inversely associated with cognitive function similar to the sensitivity results in BPRHS. Glucose was also inversely associated with cognitive function when restricting the HCHS/SOL Puerto-Rican and the total HCHS/SOL analytic sample to participants age 60 and above and restricting HCHS/SOL Puerto Ricans to participants with high adherence to the Mediterranean diet. All sensitivity analyses are presented in Supplementary Figures 1 & 2 and summarized in Supplementary Table 4.

### 3.3 Generalization analysis of metabolite associations in ARIC and meta-analysis

Figure 4 and Supplementary Table 5 summarize previous association results of metabolite associations with global cognitive function in the BPRHS, together with generalization results in the total HCHS/SOL population and the European and African American ARIC populations, and their meta-analyses with BPRHS. In general, 6 out of 10 metabolite associations had a consistent negative direction of estimated effects i.e., worse cognitive function, between BPRHS, HCHS/SOL, and ARIC, for both Europeans and African Americans, with a significant meta-analysis p-value: 5’-Methylthioadenosine, glucose, mannose, ribitol, gamma-CEHC glucuronide, mannitol/sorbitol. The first four metabolites were nominally significant in ARIC Europeans, and glucose was also nominally significant in ARIC African Americans.

**Figure 4:**
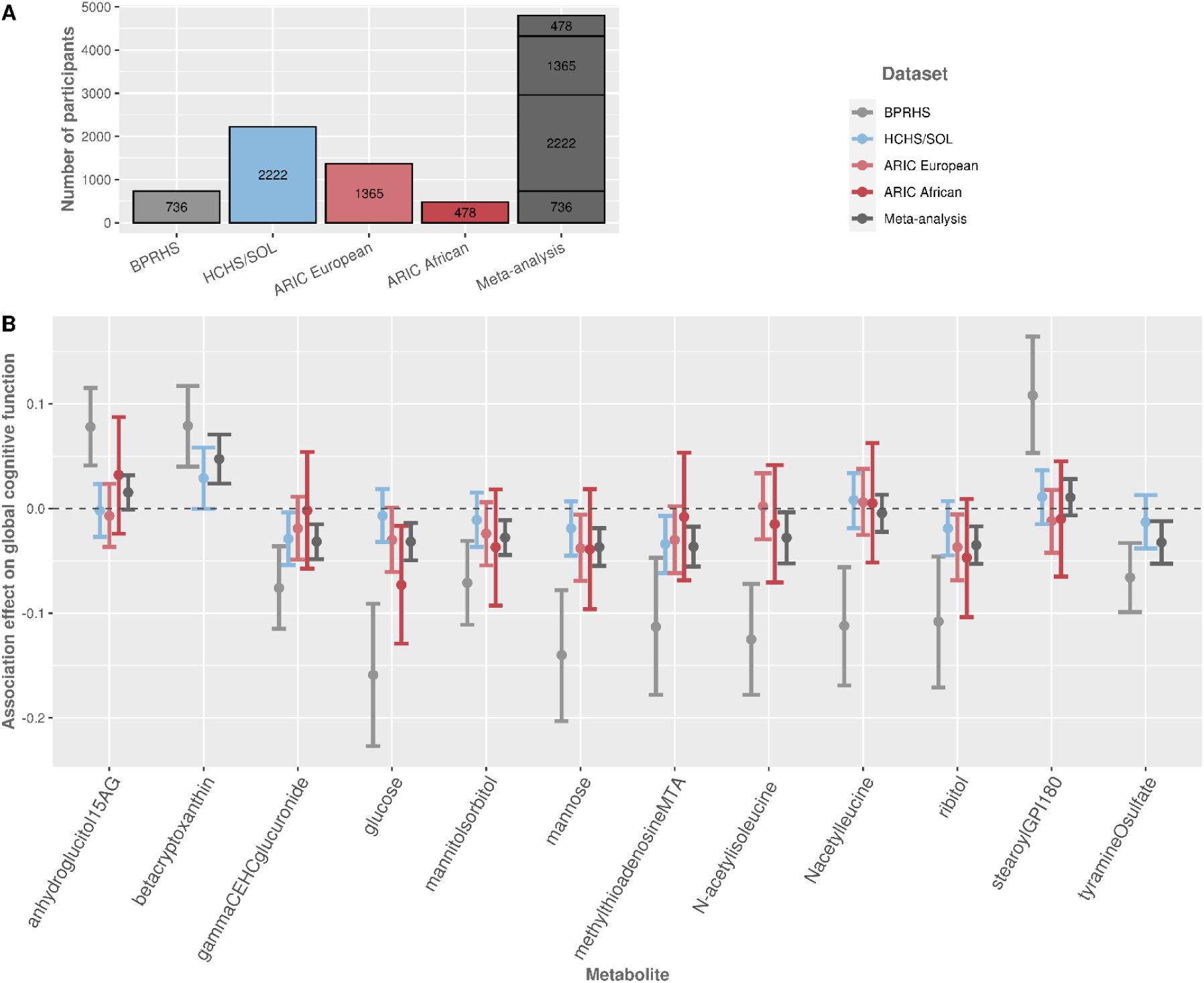
**A**. The number of participants in each study and/or combination of studies. **B**. Estimated effects and 95% confidence intervals for single metabolite associations with global cognitive function, in BPRHS (previously reported), in HCHS/SOL total analytical dataset, and ARIC Europeans and African Americans. Meta-analysis of all the groups together is presented as well (total sample sizes vary by metabolite, due to missing metabolite data in specific studies). Models are adjusted for age, sex, study center, education, BMI, smoking, *APOE*-ε4 carrier status, smoking, and Mediterranean diet score.

### 3.4 Mendelian Randomization - Single metabolites and global cognitive function

MR analyses based on GWAS of general cognitive function from Europeans (n=300,486)[28] did not support causal effects of metabolites on general cognitive function (Supplementary Table 6). MR analyses based on metabolite GWAS summary statistics from Hispanics/Latinos and cognitive performance from Europeans (n=257,841)[29] resulted in weak evidence for a potential causal effect of ribitol on cognitive performance using two variants as instrumental variables with the IVW multiplicative random-effects method (p-value= 0.03) (Supplementary Table 7). The 2 instrumental variables explained R^2^= 3.69% of the ribitol (exposure) variance. For all other metabolites, we found no evidence for causal effects on cognitive performance (Supplementary Table 7). Across metabolites, the explained variance by the instrumental variables ranged between 0.94-12.43%.

### 3.4 Assessment of dietary contributions to the association between metabolites and cognitive function

Diet scores and metabolites’ association results in HCHS/SOL are presented in Supplementary Table 8. Mediterranean diet and its dietary scores components presented significant associations with many of the metabolites, with the strongest positive association between the fruit score and beta-Cryptoxanthin (beta=0.38, p-value=2.69E-26). Bi-directional MR results for single-food intake scores and cognitive performance are presented in Supplementary Tables 9 & 10. Suggestive evidence for causality was detected for both directions, with stronger significant associations for a causal effect of cognitive performance on single-food intake scores. Further details are summarized in the supplemental data.

## 4. Discussion

In this study, we replicated previously reported metabolite associations with general cognitive function in older U.S. Puerto-Rican adults and generalized them to other population groups, including Hispanics/Latinos in the U.S., European Americans, and African Americans. Higher blood plasma levels of six metabolites were associated with lower general cognitive function in all populations: gamma-CEHC glucuronide, 5’-Methylthioadenosine, glucose, mannose, ribitol, mannitol/sorbitol. In addition, beta-Cryptoxanthin and Tyramine-O-sulfate, were associated with higher and lower global cognitive function in the HCHS/SOL Puerto-Ricans respectively, and beta-Cryptoxanthin association also generalized to the total analytic HCHS/SOL dataset (data for these two metabolites was not available in ARIC). Assessing causal association using MR for metabolites and cognitive performance suggested that the association with ribitol is potentially causal, but the evidence is weak given multiple testing (unadjusted p-value=0.03). All metabolites with replicated and/or generalized association with global cognitive function, other than mannitol/sorbitol, were associated with the Mediterranean diet and/or its component food groups/nutrients scores.

The six generalized metabolites that were negatively associated with cognitive function across race/ethnicities are positively correlated with each other in the HCHS/SOL, as shown in Figure 3. Four of them are sugars or sugar derivatives: glucose, mannose, ribitol, and mannitol/sorbitol. Glucose is the main source of energy of the human brain, and glucose metabolism monitoring is critical for brain physiology[30]. In the secondary analysis excluding individuals with Diabetes, there was no negative association between glucose and cognitive function. Dysregulation of blood sugars was previously linked to cognitive impairment; however, clinical studies have not demonstrated a beneficial effect of glucose regulation on cognitive outcomes[31]. This is compatible with the mostly null results from MR causality assessment between the metabolites and cognitive function. Nevertheless, MR analysis did suggest that ribitol may have a causal effect on cognitive performance using strong genetic instrumental variables (R^2^= 3.69%). Ribitol is pentose alcohol formed by the reduction of ribose. A recent study in China of serum metabolomic profiling in patients with Alzheimer’s disease and amnestic mild cognitive impairment included ribitol as one of the metabolites discriminating between the disease groups and the normal healthy control group[32]. Ribitol concentrations were lower in the disease groups and ribitol was positively correlated with cognitive assessment scores. The incompatibility of these results with our results could be due to genetic and environmental differences, potentially influencing the metabolite levels and cognitive function differently, or the published association may be a false positive association. Further studies are needed to elucidate the effect of ribitol on cognitive function across race/ethnicities.

Beta-Cryptoxanthin, which was positively associated with cognition in both Puerto Ricans and the total HCHS/SOL population, is an antioxidant, pre-vitamin A carotenoid found in fruits and vegetables. It was previously suggested it may be involved with cancer prevention and decreased risk of insulin resistance and liver dysfunction[33]. MR analysis, with limited power, using one instrumental variable which explained <1% of the metabolite variance, did not support a causal effect of beta-Cryptoxanthin on cognitive performance. Beta-Cryptoxanthin was associated with multiple diet scores in HCHS/SOL, with fruit score being the most prominent association (beta=0.42, p-value=4.56E-30). This supports the important role of a healthy diet in cognitive health.

Based on our analysis, the evidence for a causal association between the specific metabolites and cognitive function is weak. However, it is possible that these metabolites are markers of an underlying causal relationship between diet and cognitive function. For example, dietary factors may cause changes in serum metabolite levels and influence cognitive function in parallel processes, resulting in metabolite-cognitive function associations. In the other direction, cognitive function may influence the choices of dietary intake (via its association with other socioeconomic factors), which induces metabolite levels, again, resulting in metabolite-cognitive function associations. Observational studies suggest a beneficial effect of the Mediterranean diet on older adults’ global cognition[34]. A recent longitudinal prevention trial showed that better adherence to an energy-restricted Mediterranean diet is associated with improved global cognition three years later[9]. However, our analysis of bi-directional MR for single-food intake scores and cognitive performance mainly supports the reverse association, where cognitive function influences nutrition. The causal effect of cognitive performance on alcohol consumption was the strongest effect of all single food-intake scores.

Overall, we demonstrated high validity and generalizability of several metabolite associations with global cognitive function across diverse race/ethnicities, specifically metabolites related to sugars. Other metabolites may be specific to race/ethnic groups. The associated metabolites also highlight the importance of a healthy diet, specifically fruit and vegetable consumption, in the association with global cognitive function in adults. Future work should study the usefulness of metabolomics biomarkers of global cognitive function across diverse race/ethnicities, preferably in study designs allowing causal inference. Our study has a few limitations. The global cognitive function score was calculated based on a different set of cognitive tests in each of the datasets, BPRHS, SOL, and ARIC. Despite this limitation, our results showed strong consistency which strengthens the validity of the findings. In addition, our study design is cross-sectional, therefore causal inference is limited. We attempted causal inference by using MR analyses that overcome unmeasured confounding. Our results showed weak causal effects, but this may be due to the differences in measurements of studied exposures and/or outcomes. Future studies are needed to evaluate metabolite associations and evaluate causal associations more precisely.

## Data Availability

https://sites.cscc.unc.edu/aric/distribution-agreements

## 5. Acknowledgments

The authors thank the staff and participants of HCHS/SOL for their important contributions. Investigators’ website - https://sites.cscc.unc.edu/hchs/.

## 6. Funding

The Hispanic Community Health Study/Study of Latinos is a collaborative study supported by contracts from the National Heart, Lung, and Blood Institute (NHLBI) to the University of North Carolina (HHSN268201300001I / N01-HC-65233), University of Miami (HHSN268201300004I / N01-HC-65234), Albert Einstein College of Medicine (HHSN268201300002I / N01-HC-65235), University of Illinois at Chicago (HHSN268201300003I / N01-HC-65236 Northwestern Univ), and San Diego State University (HHSN268201300005I / N01-HC-65237). The following Institutes/Centers/Offices have contributed to the HCHS/SOL through a transfer of funds to the NHLBI: National Institute on Minority Health and Health Disparities, National Institute on Deafness and Other Communication Disorders, National Institute of Dental and Craniofacial Research, National Institute of Diabetes and Digestive and Kidney Diseases, National Institute of Neurological Disorders and Stroke, NIH Institution-Office of Dietary Supplements. This work was supported by the National Institute on Aging (R21AG070644, R01AG048642, RF1AG054548, RF1AG061022, and R21AG056952). Dr. González also receives additional support from P30AG062429 and P30AG059299. Support for metabolomics data was graciously provided by the JLH Foundation (Houston, Texas). The Atherosclerosis Risk in Communities study has been funded in whole or in part with Federal funds from the National Heart, Lung, and Blood Institute, National Institutes of Health, Department of Health and Human Services under contract numbers (HHSN268201700001I, HHSN268201700002I, HHSN268201700003I, HHSN268201700004I, and HHSN268201700005I). The authors thank the staff and participants of the ARIC study for their important contributions. The metabolomics research was sponsored by the National Human Genome Research Institute (3U01HG004402-02S1).

## Notes

Declarations of interests: BSK is the inventor on general metabolomics-related IP that has been licensed to Metabolon via Weill Medical College of Cornell University and for which he receives royalty payments via Weill Medical College of Cornell University. He also consults for and has a small equity interest in the company. Metabolon offers biochemical profiling services and is developing molecular diagnostic assays detecting and monitoring disease. Metabolon has no rights or proprietary access to the research results presented and/or new IP generated under these grants/studies. BSK’s interests were reviewed by the Brigham and Women’s Hospital and Partners Healthcare in accordance with their institutional policy. Accordingly, upon review, the institution determined that BSK’s financial interest in Metabolon does not create a significant financial conflict of interest (FCOI) with this research. The addition of this statement where appropriate was explicitly requested and approved by BWH.

### Competing Interest Statement

BSK is the inventor on general metabolomics-related IP that has been licensed to Metabolon via Weill Medical College of Cornell University and for which he receives royalty payments via Weill Medical College of Cornell University. He also consults for and has a small equity interest in the company. Metabolon offers biochemical profiling services and is developing molecular diagnostic assays detecting and monitoring disease. Metabolon has no rights or proprietary access to the research results presented and/or new IP generated under these grants/studies. BSKs interests were reviewed by the Brigham and Womens Hospital and Partners Healthcare in accordance with their institutional policy. Accordingly, upon review, the institution determined that BSKs financial interest in Metabolon does not create a significant financial conflict of interest (FCOI) with this research. The addition of this statement where appropriate was explicitly requested and approved by BWH.

### Author Declarations

The study was reviewed and approved by the Institutional Review Boards at all collaborating institutions.

