## Supplemental data for "Plasma metabolites associated with cognitive function across race/ethnicities affirming the importance of healthy nutrition"

### Plasma metabolites associated with cognitive function across race/ethnicities with a potential causal role for ribitol

##### A. ARIC Methods

The Atherosclerosis Risk in Communities (ARIC) study is a prospective longitudinal study of the development of atherosclerosis, and its clinical sequelae in which 15,792 individuals aged 45-64 years from four communities in the United States were enrolled at the baseline examination (1987-1989). A detailed description of the ARIC study has been reported previously[1]. Various medical information and specimens were collected at each of the eight examinations. In particular, cognitive testing for this study was performed in the entire cohort at the 2nd examination in 1990-1992. Written informed consent was provided by all study participants, and the study design and methods were approved by the institutional review boards at each of the collaborating medical institutions: University of Mississippi Medical Center Institutional Review Board (Jackson Field Center); of UNC Chapel Hill Institutional Review Board (Forsyth County Field Center); University of Minnesota Institutional Review Board (Minnesota Field Center); and Johns Hopkins University School of Public Health Institutional Review Board (Washington County Field Center). In this analysis, we use 1,365 European American and 478 African American ARIC participants who had measured metabolites in serum from their 1$^{st}$ clinic visit. We excluded participants who had (1) unknown history of definite or probable stroke or a history of physician-diagnosed stroke prior to visit 2, or (2) if they were missing any of the three cognitive tests scores or information for any covariates. Detailed descriptions of participants' health, demographic, and lifestyle characteristics are presented in Table 1.

##### B. Mendelian Randomization analysis to assess a causal effect of metabolites on global cognitive function

We performed a two-sample MR to estimate the causal effect of each of the replicated metabolites (exposures) on global cognitive function (outcome) using the 'TwoSampleMR' R package[2] (Figure 1, Step D). We first performed genome-wide association studies (GWAS) using the 'GENESIS' R package for each of the 8 metabolites (showing evidence of replication and/or generalization in HCHS/SOL and/or ARIC) in 3,930 HCHS/SOL individuals who had both genetic data and metabolite data, adjusting for age, sex, center, estimated glomerular filtration rate, log of the sample weights, first 5 PCs, and genetic analysis group. We removed genetic variants with low minor allele frequency (MAF) (<0.01), low minor allele count (MAC) (≤50), and low imputation quality (<0.6), and selected variants with P-value threshold ≤ 1e-7 as genetic instruments, for each metabolite separately. To ensure that the instruments for the exposure are independent, we used the 'ld_clump_local' function, using HCHS/SOL imputed data as a reference panel, with the following parameters: clump_kb=5000, clump_r2=0.1, clump_p=1 (e.g. for a pair of genetic variants within 5,000K base pairs and R2 correlation > 0.1, one variant will be excluded). For each of the metabolites, we computed their variance explained by their genetic instruments as the percent change in residual variance in regression of these metabolites over covariates, compared to regression of these metabolites over covariates and the genetic instruments. For this computation, we used a subset of individuals who are genetically unrelated and do not live in the same household.

Because the GWAS summary statistics used were for general cognitive function European ancestry populations, we also used summary statistics of blood metabolite GWAS conducted in 1,960 adults of European ancestry, for 5 (out of the 11) metabolites: 5-methylthioadenosine (MTA), glucose, 15-anhydroglucitol (15-AG), mannose, and ribitol[3]. After removing genetic variants with low MAF (<0.01), we selected variants with P-value threshold ≤ 1e-7 as genetic instruments, for each metabolite separately, and then used the 'clump_data' function with the European reference panel to ensure that the instruments for each exposure are independent of each other.

We used summary statistics from a GWAS of cognitive performance based on 257,841 European individuals from COGENT consortium and UK Biobank[4], and from another similar GWAS of general cognitive function based on 300,486 European individuals (ages 16-102) from the CHARGE consortium, COGENT consortium, and UK Biobank[5]. The cognitive outcomes of these GWAS were not measured in the same way for all participants within each study and were different from our global cognitive function score, therefore they are used as proxy phenotypes. After harmonizing the exposure and outcome data (matching genetic variants and alleles from exposure and outcomes GWAS), we performed the MR analysis with a range of different MR methods offered by the package: Inverse variance weighted (IVW), MR Egger, weighted median, and Wald ratio when there was only a single instrument. Our primary method was IVW with multiplicative random effects since it is recommended when using summarized data, and it allows for heterogeneity across instruments[6]. Heterogeneity and horizontal pleiotropy were also assessed using the 'TwoSampleMR' R package[2].

##### C. Association between diet scores and metabolites in HCHS/SOL

We studied the associations between the Mediterranean diet and the metabolites among the total HCHS/SOL analytic population, using generalized linear mixed models, (Figure1, Step E). In model 1, we tested the cross-sectional association between the Mediterranean diet score with each of the 8 metabolites showing evidence of replication and/or generalization in HCHS/SOL and ARIC, and in model 2 we tested the association between each of the 9 dietary scores comprising the Mediterranean diet score (together in the same model), with each of the 8 metabolites. We adjusted for age, sex, study center, education (<12,12,12< years), BMI (normal, overweight, obese), smoking (never, past, current), APOE-ε4 carrier status (dominant mode), smoking (never, past, current), and self-identified Hispanic/Latino background.

Supplementary Table 8 presents the associations between the Mediterranean diet score (model 1) and each of the 8 metabolites, and between each of the 9 diet scores (all included in model 2) with each of the 8 metabolites, in the total HCHS/SOL analytic sample (n=2307). Mediterranean diet is significantly associated with 5 of the metabolites, with the strongest positive association for betacryptoxanthin (beta=0.122, p-value=2.8E-25). Other than the Mufa_sfa score, all other 8 dietary scores, presented significant associations with at least one of the 8 metabolites. The most prominent is the fruit score, associated with four of the metabolites, showing the strongest positive association with betacryptoxanthin (beta=0.38, p-value=2.69E-26). Overall, betacryptoxanthin presented the highest number of significant associations with five out of nine dietary scores.

#### D. Bi-directional Mendelian Randomization: single-food intake scores and cognitive performance

We performed bidirectional MR using the 'TwoSampleMR' R package to estimate the causal effect of single-food intake scores (exposure) on cognitive performance (outcome), and vice-versa, the causal effect of cognitive performance (exposure) on single-food intake scores (outcome)[2] (Figure1, Step F). We utilized GWAS summary statistics of single-food intake scores, conducted in up to 449,210 Europeans from UK Biobank[7]. We selected 12 single-food intake scores related to the 9 dietary components of the Mediterranean diet: freshfruit, driedfruit, rawveg, cookedveg, beef, lambmutton, pork, poultry, processmeat, nonoilyfish, oilyfish, alcohol. Further details on single-food intake scores can be found here: http://kp4cd.org/sites/default/files/READMEs/Cole_UKB_Diet_GWAS_README.pdf. The GWAS results were previously filtered as follows: MAC ≥ 20, MAF ≥ 0.005, INFO score ≥ 0.60.

When using single-food intake score as an exposure, for each score we selected variants with a P-value threshold <5e-8 as genetic instruments, and then used the 'clump_data' function with the European reference panel to ensure that the instruments for the exposure are independent. We used summary statistics from a GWAS of cognitive performance based on 257,841 European individuals from COGENT consortia, and UK Biobank[4]. When using cognitive performance as an exposure, we selected variants with a P-value threshold <1e-7 as genetic instruments and then used the 'clump_data' function with the European reference panel to ensure that the instruments for the exposure are independent.

After harmonizing the exposure and outcome data, we performed the MR analysis with a range of different MR methods offered by the package: Inverse variance weighted (IVW), MR Egger, weighted median, and Wald ratio when there was only a single instrument. Our primary method was IVW with multiplicative random effects since it is recommended when using summarized data, and it allows for heterogeneity across instruments[6].

Supplementary Table 9 presents the results of the MR analyses with suggestive evidence of a causal negative association between alcohol (using 75 instrumental variables, p-value=5.34E-04) and dried fruits (using 33 instrumental variables, p-value=0.01) with cognitive performance. Supplementary Table 10 presents the inverse direction of causality, between cognitive performance and single-food intake scores (using 162 instrumental variables) with stronger significant negative associations with alcohol (p-value=2.06E-17) and dried fruits consumption (p-value=1.16E-07), as well as significant associations with other single-food intake scores, including beef, cooked vegetables, lamb mutton, non-oily fish, poultry, and raw vegetables, all presenting a positive association, other than lamb mutton.

**Supplementary Figure 1:** Sensitivity analyses of single metabolites associations with global cognitive function in HCHS/SOL Puerto-Ricans.

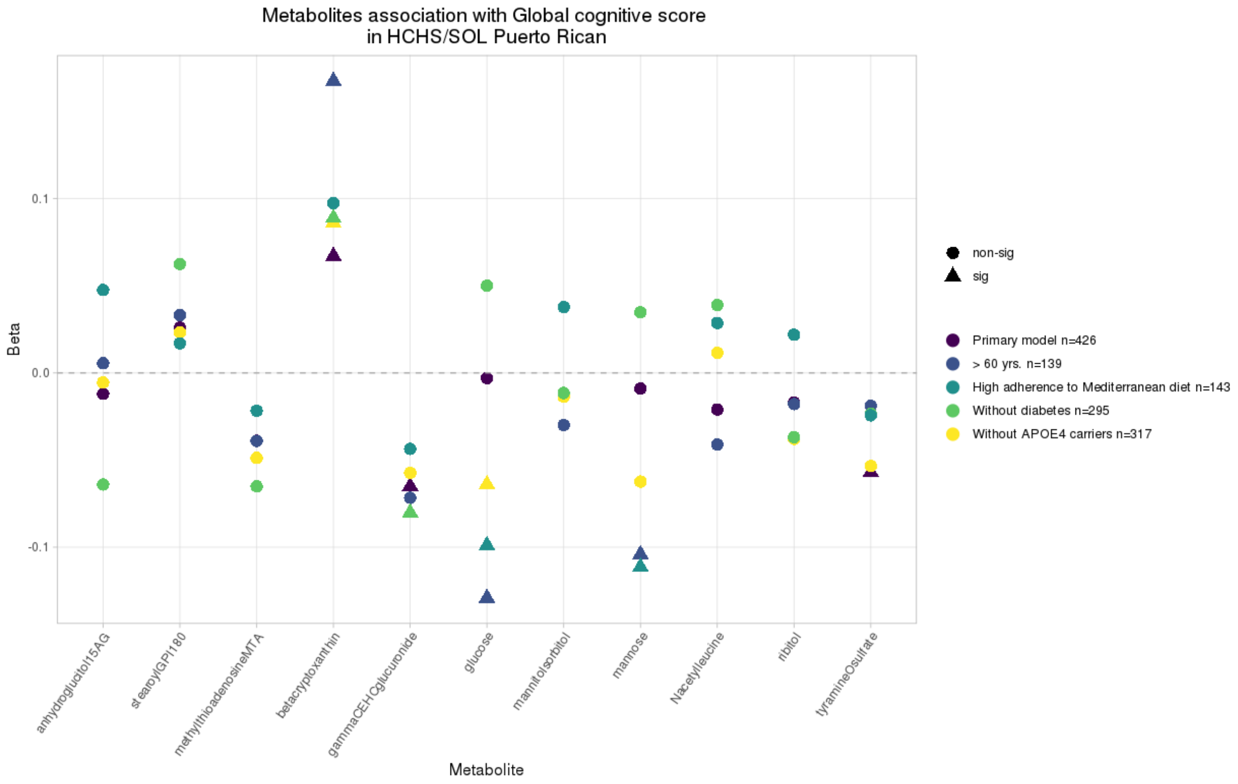

**Supplementary Figure 2:** Sensitivity analyses of single metabolites associations with global cognitive function in the total HCHS/SOL analytic sample.

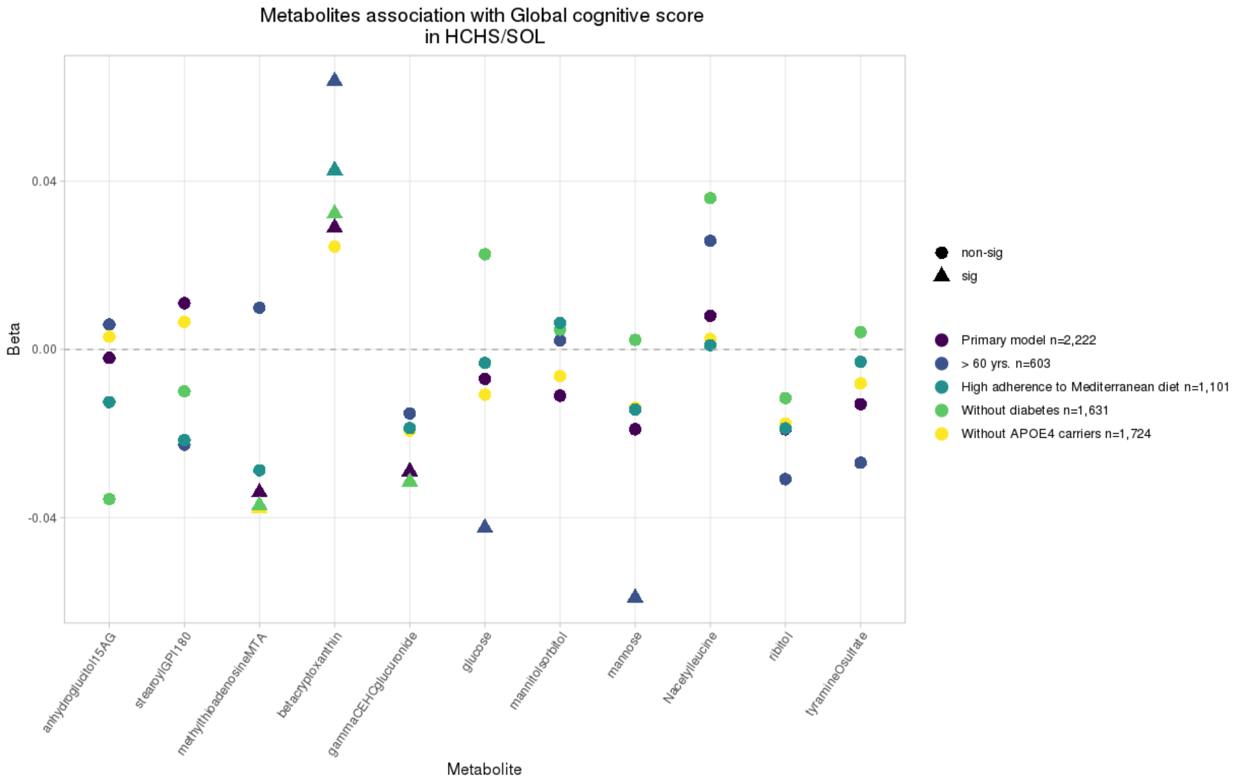

Supplementary Table 1: Previously reported single metabolites associated with global cognitive function in the BPRHS and their matching names in HCHS/SOL and ARIC studies.

| | Metabolite name | | |
|---|---|---|---|
| **HMDB** | **BPRHS** | **HCHS/SOL** | **ARIC** |
| HMDB02712 | 1,5-anhydroglucitol (1,5-AG) | 1,5-anhydroglucitol (1,5-AG) | 1,5-anhydroglucitol (1,5-AG) |
| HMDB33844 | beta- cryptoxanthin | beta-cryptoxanthin | NA |
| HMDB61696 | 1-stearoyl-GPI (18:0) | 1-stearoyl-GPI (18:0) | 1-stearoylglycerophosphoinositol |
| HMDB00122 | glucose | glucose | glucose |
| HMDB06409 | tyramine O-sulfate Amino Acid | tyramine O-sulfate | NA |
| HMDB00854 | formiminoglutamate Amino Acid | NA | NA |
| HMDB00508 | ribitol | ribitol | ribitol |
| HMDB01173 | 5-methylthioadenosine (MTA) | 5-methylthioadenosine (MTA) | 5-methylthioadenosine (MTA) |
| HMDB61684 | N-acetylisoleucine Amino Acid | NA | N-acetylisoleucine |
| HMDB00169 | mannose | mannose | mannose |
| HMDB11756 | N-acetylleucine | N-acetylleucine | N-acetylleucine |
| HMDB00247 | mannitol/sorbitol | mannitol/sorbitol | sorbitol |
| - | gamma-CEHC glucuronide* | gamma-CEHC glucuronide* | gamma-CEHC glucuronide* |

**Abbreviations:** *HMDB* , The Human Metabolome Database; *BPRHS* , Boston Puerto-Rican Health Study; *HCHS/SOL* , Hispanic Community Health Study/ Study of Latinos; *ARIC* , The Atherosclerosis Risk in Communities Study.

Supplementary Table 2: Previously reported metabolites included in the MRS (n=51) associated with global cognitive function in the BPRHS and their matching names in HCHS/SOL (n=36).

| | Metabolite name | |
|---|---|---|
| HMDB | BPRHS | HCHS/SOL |
| HMDB01999 | eicosapentaenoate | eicosapentaenoate (EPA; 20:5n3) |
| HMDB02329 | oxalate (ethanedioate) | oxalate (ethanedioate) |
| HMDB07218 | oleoyl-oleoyl-glycerol | NA |
| HMDB00574 | Cysteine Amino Acid Methionine, Cysteine, SAM | cysteine |
| HMDB04705 | 12,13-DiHOME Lipid Fatty Acid, Dihydroxy | 12,13-DiHOME |
| HMDB02823 | docosatrienoate (22 : 3n3) Lipid Polyunsaturated Fatty Acid | NA |
| HMDB33844 | beta-cryptoxanthin Cofactors and Vitamins Vitamin A Metabolism | beta-cryptoxanthin |
| HMDB08141 | 1-linoleoyl-2-linolenoyl-GPC | 1-linoleoyl-2-linolenoyl-GPC (18:2/18:3)* |
| HMDB00345 | 3-hydroxyadipate* Lipid Fatty Acid, Dicarboxylate | NA |
| HMDB00017 | Pyridoxate Cofactors and Vitamins Vitamin B6 Metabolism | pyridoxate |
| HMDB00661 | glutarate (C5-DC) Lipid Fatty Acid, Dicarboxylate | glutarate (pentanedioate) |
| HMDB61115 | tryptophan betaine Amino Acid Tryptophan Metabolism | tryptophan betaine |
| HMDB00202 | methylmalo-te (MMA) Lipid Fatty Acid Metabolism (also | NA |
| HMDB61684 | N-acetylisoleucine Amino Acid Leucine, Isoleucine and | NA |
| HMDB00190 | lactate Carbohydrate Glycolysis, Gluconeogenesis, | lactate |
| HMDB06409 | tyramine O-sulfate Amino Acid Tyrosine Metabolism | tyramine O-sulfate |
| HMDB00063 | Cortisol Lipid Corticosteroids | cortisol |
| HMDB07257 | linoleoyl-arachidonoylglycerol (18 : 2/20 : 4) | linoleoyl-arachidonoyl-glycerol (18:2/20:4) [1]* |
| HMDB03464 | 4-guanidinobutanoate Amino Acid Guanidino and Acetamido | 4-guanidinobutanoate |
| HMDB02802 | cortisone Lipid Corticosteroids | cortisone |
| HMDB00258 | sucrose | sucrose |
| HMDB07228 | oleoyl-arachidonoyl-glycerol | oleoyl-arachidonoyl-glycerol (18:1/20:4) [1]* |
| HMDB00126 | glycerol 3-phosphate | glycerol 3-phosphate |
| HMDB09069 | 1-oleoyl-2-arachidonoyl-GPE | NA |
| HMDB00012 | 2’-deoxyuridine | 2'-deoxyuridine |
| HMDB11745 | N-acetylmethionine | N-acetylmethionine |
| HMDB00522 | 3-methylglutaco-te | 3-methylglutaconate |
| HMDB01547 | corticosterone | NA |
| HMDB11756 | N-acetylleucine | N-acetylleucine |
| HMDB00631 | glycodeoxycholate | glycodeoxycholate |
| HMDB00122 | glucose | glucose |
| HMDB00754 | beta-hydroxyisovalerate | beta-hydroxyisovalerate |
| HMDB02820 | 1-methyl-4-imidazoleacetate Amino Acid | 1-methylimidazoleacetate |
| HMDB09789 | 1-palmitoyl-2-arachidonoyl- | 1-palmitoyl-2-arachidonoyl-GPI (16:0/20:4)* |
| HMDB00669 | 2-hydroxyphenylacetate | 2-hydroxyphenylacetate |
| HMDB04089 | N-formylanthranilic acid | N-formylanthranilic acid |
| HMDB10395 | 1-arachidonoyl-GPC | 1-arachidonoyl-GPC (20:4n6)* |
| - | 4-methoxyphenol sulfate | NA |
| - | N-stearoylserine | NA |
| - | 3-hydroxyhexanoate | 3-hydroxyhexanoate |
| - | lignoceroyl sphingomyelin (d18 : 1/24 : 0) | lignoceroyl sphingomyelin (d18:1/24:0) |
| - | 1-stearoyl-2-oleoyl-GPG (18 : 0/18 : 1) | NA |
| - | sphingomyelin (d18 : 2/16 : 0,d18 : 1/16 : 1) | NA |
| - | acetylspermidine | NA |
| - | arabote/xylote | NA |
| - | glycosyl-N-(2-hydroxynervonoyl)-sphingosine(d18 : 1/24 : 1(2OH)) | glycosyl-N-(2-hydroxynervonoyl)-sphingosine (d18:1/24:1(2OH))* |
| - | 3-methylcytidine | NA |
| - | gamma-glutamylcitrulline | NA |
| - | sphingomyelin (d18 : 2/18 : 1) | sphingomyelin (d18:2/18:1)* |
| - | acisoga | acisoga |
| - | cis-4-decenoylcarnitine (C10 : 1) | cis-4-decenoylcarnitine (C10:1) |

**Abbreviations:** *MRS* , Metabolites risk score; HMDB, The Human Metabolome Database; BPRHS, Boston Puerto-Rican Health Study; HCHS/SOL, Hispanic Community Health Study/ Study of Latinos.

Supplementary Table 3: Metabolite associations with global cognitive function in BPRHS, HCHS/SOL, and meta-analyzed. Models are adjusted for: age, sex, study center, education, BMI, smoking, APOE-ε4 carrier status, smoking, and Mediterranean diet score. In the total HCHS/SOL analytic sample model adjustment was done also for self-identified Hispanic/Latino background. Effect sizes and SEs were estimated based on the generalized mixed-models analysis approach.

| | | BPRHS n=736 | | | | | | HCHS/SOL | | | | Meta_analysis | | | | | | |
|---|---|---|---|---|---|---|---|---|---|---|---|---|---|---|---|---|---|---|
| SOL population | Metabolite | Beta | CI low | CI high | P-value | Adjusted P value | SE | Beta | SE | P-value | P-value1 sided | Beta | se | Z | P-value Z | direction | Q | pval.Q |
| Puerto-Rican n=426 | anhydroglucitol15AG | 0.078 | 0.041 | 0.115 | 3.78E-05 | 5.87E-03 | 0.019 | -0.012 | 0.031 | 7.00E-01 | 6.50E-01 | 0.054 | 0.016 | 3.315 | 9.16E-04 | +- | 6.139 | 1.32E-02 |
| | stearoylGPI180 | 0.108 | 0.053 | 0.164 | 1.19E-04 | 9.21E-03 | 0.028 | 0.026 | 0.031 | 4.13E-01 | 2.07E-01 | 0.071 | 0.021 | 3.416 | 6.36E-04 | ++ | 3.845 | 4.99E-02 |
| | methylthioadenosineMTA | -0.113 | -0.178 | -0.047 | 7.81E-04 | 3.73E-02 | 0.034 | -0.039 | 0.031 | 2.18E-01 | 1.09E-01 | -0.073 | 0.023 | -3.202 | 1.37E-03 | -- | 2.617 | 1.06E-01 |
| | betacryptoxanthin | 0.079 | 0.040 | 0.117 | 6.92E-05 | 8.59E-03 | 0.020 | 0.067 | 0.035 | 5.72E-02 | 2.86E-02 | 0.076 | 0.017 | 4.405 | 1.06E-05 | ++ | 0.089 | 7.66E-01 |
| | gammaCEHCglucuronide | -0.076 | -0.115 | -0.036 | 1.71E-04 | 1.18E-02 | 0.020 | -0.065 | 0.031 | 3.79E-02 | 1.90E-02 | -0.073 | 0.017 | -4.293 | 1.76E-05 | -- | 0.088 | 7.66E-01 |
| | glucose | -0.159 | -0.227 | -0.091 | 6.06E-06 | 1.88E-03 | 0.035 | -0.003 | 0.030 | 9.10E-01 | 4.55E-01 | -0.069 | 0.023 | -3.013 | 2.58E-03 | -- | 11.398 | 7.35E-04 |
| | mannitolsorbitol | -0.071 | -0.111 | -0.031 | 5.59E-04 | 3.16E-02 | 0.021 | -0.012 | 0.032 | 7.02E-01 | 3.51E-01 | -0.054 | 0.017 | -3.105 | 1.90E-03 | -- | 2.405 | 1.21E-01 |
| | mannose | -0.140 | -0.203 | -0.078 | 1.17E-05 | 2.41E-03 | 0.032 | -0.009 | 0.032 | 7.72E-01 | 3.86E-01 | -0.075 | 0.023 | -3.301 | 9.64E-04 | -- | 8.395 | 3.76E-03 |
| | Nacetylleucine | -0.112 | -0.169 | -0.056 | 1.11E-04 | 9.21E-03 | 0.029 | -0.021 | 0.029 | 4.63E-01 | 2.32E-01 | -0.067 | 0.020 | -3.246 | 1.17E-03 | -- | 4.927 | 2.64E-02 |
| | ribitol | -0.108 | -0.171 | -0.046 | 7.12E-04 | 3.68E-02 | 0.032 | -0.017 | 0.029 | 5.65E-01 | 2.83E-01 | -0.058 | 0.021 | -2.710 | 6.72E-03 | -- | 4.455 | 3.48E-02 |
| | tyramineOsulfate | -0.066 | -0.099 | -0.033 | 1.11E-04 | 9.21E-03 | 0.017 | -0.057 | 0.031 | 6.88E-02 | 3.44E-02 | -0.064 | 0.015 | -4.273 | 1.93E-05 | -- | 0.065 | 7.99E-01 |
| Total n=2,222 | anhydroglucitol15AG | 0.078 | 0.041 | 0.115 | 3.78E-05 | 5.87E-03 | 0.019 | -0.002 | 0.013 | 8.90E-01 | 5.55E-01 | 0.024 | 0.011 | 2.206 | 2.74E-02 | +- | 12.137 | 4.94E-04 |
| | stearoylGPI180 | 0.108 | 0.053 | 0.164 | 1.19E-04 | 9.21E-03 | 0.028 | 0.011 | 0.013 | 4.10E-01 | 2.05E-01 | 0.028 | 0.012 | 2.385 | 1.71E-02 | ++ | 9.835 | 1.71E-03 |
| | methylthioadenosineMTA | -0.113 | -0.178 | -0.047 | 7.81E-04 | 3.73E-02 | 0.034 | -0.034 | 0.014 | 1.39E-02 | 6.95E-03 | -0.046 | 0.013 | -3.533 | 4.11E-04 | -- | 4.702 | 3.01E-02 |
| | betacryptoxanthin | 0.079 | 0.040 | 0.117 | 6.92E-05 | 8.59E-03 | 0.020 | 0.029 | 0.015 | 5.35E-02 | 2.68E-02 | 0.047 | 0.012 | 3.941 | 8.11E-05 | ++ | 4.038 | 4.45E-02 |
| | gammaCEHCglucuronide | -0.076 | -0.115 | -0.036 | 1.71E-04 | 1.18E-02 | 0.020 | -0.029 | 0.013 | 2.40E-02 | 1.20E-02 | -0.043 | 0.011 | -3.909 | 9.27E-05 | -- | 3.822 | 5.06E-02 |
| | glucose | -0.159 | -0.227 | -0.091 | 6.06E-06 | 1.88E-03 | 0.035 | -0.007 | 0.013 | 6.05E-01 | 3.03E-01 | -0.025 | 0.012 | -2.075 | 3.80E-02 | -- | 16.455 | 4.98E-05 |
| | mannitolsorbitol | -0.071 | -0.111 | -0.031 | 5.59E-04 | 3.16E-02 | 0.021 | -0.011 | 0.013 | 4.16E-01 | 2.08E-01 | -0.028 | 0.011 | -2.559 | 1.05E-02 | -- | 6.078 | 1.37E-02 |
| | mannose | -0.140 | -0.203 | -0.078 | 1.17E-05 | 2.41E-03 | 0.032 | -0.019 | 0.013 | 1.49E-01 | 7.45E-02 | -0.036 | 0.012 | -3.006 | 2.65E-03 | -- | 12.311 | 4.50E-04 |
| | Nacetylleucine | -0.112 | -0.169 | -0.056 | 1.11E-04 | 9.21E-03 | 0.029 | 0.008 | 0.013 | 5.73E-01 | 7.14E-01 | -0.012 | 0.012 | -1.021 | 3.07E-01 | -+ | 14.277 | 1.58E-04 |
| | ribitol | -0.108 | -0.171 | -0.046 | 7.12E-04 | 3.68E-02 | 0.032 | -0.019 | 0.013 | 1.53E-01 | 7.65E-02 | -0.032 | 0.012 | -2.631 | 8.52E-03 | -- | 6.673 | 9.79E-03 |
| | tyramineOsulfate | -0.066 | -0.099 | -0.033 | 1.11E-04 | 9.21E-03 | 0.017 | -0.013 | 0.013 | 3.26E-01 | 1.63E-01 | -0.032 | 0.010 | -3.137 | 1.71E-03 | -- | 6.099 | 1.35E-02 |

**Abbreviations:** *HCHS/SOL* , Hispanic Community Health Study/ Study of Latinos; BPRHS, Boston Puerto-Rican Health Study; CI*,* Confidence Interval; SE, standard error.

Supplementary Table 4: Sensitivity analyses of metabolite associations with global cognitive function in HCHS/SOL Puerto-Ricans and in the total HCHS/SOL analytic sample. Models are adjusted for: age, sex, study center, education, BMI, smoking, APOE-ε4 carrier status, smoking, and Mediterranean diet score. In the total HCHS/SOL analytic sample model adjustment was done also for self-identified Hispanic/Latino background. Effect sizes and SEs were estimated based on the mixed-models analysis approach.

| | | HCHS/SOL Puerto-Ricans | | | | HCHS/SOL Total | | | |
|---|---|---|---|---|---|---|---|---|---|
| **Model** | **Metabolites** | **Beta** | **SE** | **P-value** | **P-value1 sided** | **Beta** | **SE** | **P-value** | **P-value1 sided** |
| Without APOE4 carriers Puerto Rican n=319 Total n=1,738 | anhydroglucitol15AG | -0.005 | 0.038 | 8.85E-01 | 5.57E-01 | 0.003 | 0.014 | 8.34E-01 | 4.17E-01 |
| | stearoylGPI180 | 0.023 | 0.038 | 5.43E-01 | 2.71E-01 | 0.007 | 0.015 | 6.61E-01 | 3.30E-01 |
| | methylthioadenosineMTA | -0.049 | 0.038 | 2.04E-01 | 1.02E-01 | -0.038 | 0.016 | 1.54E-02 | 7.71E-03 |
| | betacryptoxanthin | 0.086 | 0.042 | 4.24E-02 | 2.12E-02 | 0.024 | 0.017 | 1.50E-01 | 7.50E-02 |
| | gammaCEHCglucuronide | -0.057 | 0.040 | 1.46E-01 | 7.32E-02 | -0.019 | 0.015 | 1.88E-01 | 9.39E-02 |
| | glucose | -0.064 | 0.036 | 7.52E-02 | 3.76E-02 | -0.011 | 0.015 | 4.63E-01 | 2.31E-01 |
| | mannitolsorbitol | -0.014 | 0.041 | 7.38E-01 | 3.69E-01 | -0.006 | 0.015 | 6.71E-01 | 3.36E-01 |
| | mannose | -0.062 | 0.038 | 1.02E-01 | 5.09E-02 | -0.014 | 0.015 | 3.52E-01 | 1.76E-01 |
| | Nacetylleucine | 0.012 | 0.035 | 7.39E-01 | 6.31E-01 | 0.003 | 0.015 | 8.70E-01 | 5.65E-01 |
| | ribitol | -0.038 | 0.035 | 2.84E-01 | 1.42E-01 | -0.018 | 0.015 | 2.33E-01 | 1.17E-01 |
| | tyramineOsulfate | -0.053 | 0.038 | 1.64E-01 | 8.21E-02 | -0.008 | 0.015 | 5.80E-01 | 2.90E-01 |
| Age >60 yrs. Puerto Rican n=141 Total n=609 | anhydroglucitol15AG | 0.006 | 0.049 | 9.10E-01 | 4.55E-01 | 0.006 | 0.024 | 8.05E-01 | 4.02E-01 |
| | stearoylGPI180 | 0.033 | 0.053 | 5.29E-01 | 2.64E-01 | -0.023 | 0.026 | 3.84E-01 | 8.08E-01 |
| | methylthioadenosineMTA | -0.039 | 0.053 | 4.61E-01 | 2.31E-01 | 0.010 | 0.026 | 7.05E-01 | 6.47E-01 |
| | betacryptoxanthin | 0.167 | 0.062 | 6.91E-03 | 3.46E-03 | 0.064 | 0.028 | 2.44E-02 | 1.22E-02 |
| | gammaCEHCglucuronide | -0.072 | 0.053 | 1.74E-01 | 8.72E-02 | -0.015 | 0.024 | 5.19E-01 | 2.60E-01 |
| | glucose | -0.129 | 0.049 | 8.03E-03 | 4.01E-03 | -0.042 | 0.025 | 8.35E-02 | 4.18E-02 |
| | mannitolsorbitol | -0.030 | 0.065 | 6.43E-01 | 3.21E-01 | 0.002 | 0.027 | 9.37E-01 | 5.32E-01 |
| | mannose | -0.104 | 0.051 | 4.13E-02 | 2.07E-02 | -0.059 | 0.025 | 1.98E-02 | 9.90E-03 |
| | Nacetylleucine | -0.041 | 0.049 | 4.01E-01 | 2.00E-01 | 0.026 | 0.026 | 3.29E-01 | 8.36E-01 |
| | ribitol | -0.018 | 0.064 | 7.80E-01 | 3.90E-01 | -0.031 | 0.027 | 2.51E-01 | 1.25E-01 |
| | tyramineOsulfate | -0.019 | 0.057 | 7.41E-01 | 3.71E-01 | -0.027 | 0.027 | 3.19E-01 | 1.60E-01 |

| | | | | | | | | | |
|---|---|---|---|---|---|---|---|---|---|
| Without diabetes Puerto Rican n=296 Total n=1,641 | anhydroglucitol15AG | -0.064 | 0.044 | 1.43E-01 | 9.29E-01 | -0.036 | 0.018 | 4.66E-02 | 9.77E-01 |
| | stearoylGPI180 | 0.063 | 0.044 | 1.52E-01 | 7.58E-02 | -0.010 | 0.017 | 5.49E-01 | 7.26E-01 |
| | methylthioadenosineMTA | -0.065 | 0.042 | 1.20E-01 | 6.00E-02 | -0.037 | 0.017 | 2.84E-02 | 1.42E-02 |
| | betacryptoxanthin | 0.089 | 0.044 | 4.45E-02 | 2.23E-02 | 0.032 | 0.018 | 6.54E-02 | 3.27E-02 |
| | gammaCEHCglucuronide | -0.080 | 0.042 | 5.61E-02 | 2.81E-02 | -0.031 | 0.016 | 4.35E-02 | 2.18E-02 |
| | glucose | 0.050 | 0.047 | 2.92E-01 | 8.54E-01 | 0.023 | 0.019 | 2.34E-01 | 8.83E-01 |
| | mannitolsorbitol | -0.012 | 0.041 | 7.76E-01 | 3.88E-01 | 0.005 | 0.016 | 7.62E-01 | 6.19E-01 |
| | mannose | 0.035 | 0.049 | 4.77E-01 | 7.62E-01 | 0.002 | 0.019 | 9.03E-01 | 5.48E-01 |
| | Nacetylleucine | 0.039 | 0.037 | 2.88E-01 | 8.56E-01 | 0.036 | 0.016 | 2.46E-02 | 9.88E-01 |
| | ribitol | -0.037 | 0.038 | 3.30E-01 | 1.65E-01 | -0.012 | 0.016 | 4.64E-01 | 2.32E-01 |
| | tyramineOsulfate | -0.023 | 0.041 | 5.67E-01 | 2.84E-01 | 0.004 | 0.015 | 7.90E-01 | 6.05E-01 |
| High adherence to Mediterranean diet Puerto Rican n=144 Total n=1,108 | anhydroglucitol15AG | 0.048 | 0.066 | 4.72E-01 | 2.36E-01 | -0.013 | 0.018 | 4.97E-01 | 7.51E-01 |
| | stearoylGPI180 | 0.017 | 0.062 | 7.82E-01 | 3.91E-01 | -0.022 | 0.018 | 2.32E-01 | 8.84E-01 |
| | methylthioadenosineMTA | -0.022 | 0.069 | 7.53E-01 | 3.77E-01 | -0.029 | 0.020 | 1.53E-01 | 7.66E-02 |
| | betacryptoxanthin | 0.097 | 0.067 | 1.45E-01 | 7.24E-02 | 0.043 | 0.021 | 3.87E-02 | 1.93E-02 |
| | gammaCEHCglucuronide | -0.044 | 0.061 | 4.74E-01 | 2.37E-01 | -0.019 | 0.018 | 2.87E-01 | 1.43E-01 |
| | glucose | -0.099 | 0.056 | 7.59E-02 | 3.80E-02 | -0.003 | 0.018 | 8.57E-01 | 4.28E-01 |
| | mannitolsorbitol | 0.038 | 0.066 | 5.69E-01 | 7.16E-01 | 0.006 | 0.019 | 7.32E-01 | 6.34E-01 |
| | mannose | -0.111 | 0.061 | 6.58E-02 | 3.29E-02 | -0.014 | 0.018 | 4.15E-01 | 2.07E-01 |
| | Nacetylleucine | 0.029 | 0.052 | 5.84E-01 | 7.08E-01 | 0.001 | 0.019 | 9.56E-01 | 5.22E-01 |
| | ribitol | 0.022 | 0.055 | 6.91E-01 | 6.55E-01 | -0.019 | 0.019 | 3.14E-01 | 1.57E-01 |
| | tyramineOsulfate | -0.024 | 0.057 | 6.69E-01 | 3.35E-01 | -0.003 | 0.018 | 8.70E-01 | 4.35E-01 |

**Abbreviations:** *HCHS/SOL*, Hispanic Community Health Study/ Study of Latinos; *SE*, standard error.

Supplementary Table 5: Single metabolite associations with global cognitive function in BPRHS, SOL, ARIC, and meta-analyzed. Models are adjusted for: age, sex, study center, education, BMI, smoking, APOE-ε4 carrier status, smoking, Mediterranean diet score, and self-identified Hispanic/Latino background. Effect sizes and SEs were estimated based on the mixed-models analysis approach.

| | BPRHS n=736 | | | | | | SOL n=2,222 | | | | ARIC_Eur n=1,365 | | | | ARIC_Afr n=478 | | | | Meta_analysis | | | | | | | | |
|---|---|---|---|---|---|---|---|---|---|---|---|---|---|---|---|---|---|---|---|---|---|---|---|---|---|---|---|
| Metabolite | Beta | CI low | CI high | P-value | Adjusted P-value | SE | Beta | SE | P-value | P.value 1 sided | Beta | SE | P-value | P-value 1 sided | Beta | SE | P-value | P.value 1 sided | Beta | SE | Z | P-value Z | direction | Q | pval.Q | Number of strata | N total |
| anhydroglucitol15AG | 0.078 | 0.041 | 0.115 | 3.78E-05 | 5.87E-03 | 0.019 | -0.002 | 0.013 | 8.90E-01 | 5.55E-01 | -0.007 | 0.015 | 6.67E-01 | 6.67E-01 | 0.032 | 0.028 | 2.67E-01 | 1.33E-01 | 0.0153 | 0.0084 | 1.824929108 | 6.80E-02 | +--+ | 15.09 | 1.74E-03 | 4 | 4,801 |
| stearoylGPI180 | 0.108 | 0.053 | 0.164 | 1.19E-04 | 9.21E-03 | 0.028 | 0.011 | 0.013 | 4.10E-01 | 2.05E-01 | -0.012 | 0.015 | 4.18E-01 | 7.91E-01 | -0.010 | 0.028 | 7.23E-01 | 6.39E-01 | 0.0107 | 0.0089 | 1.21 | 2.26E-01 | ++-- | 14.84 | 1.96E-03 | 4 | 4,801 |
| methylthioadenosineMTA | -0.113 | -0.178 | -0.047 | 7.81E-04 | 3.73E-02 | 0.034 | -0.034 | 0.014 | 1.39E-02 | 6.95E-03 | -0.030 | 0.016 | 6.82E-02 | 3.41E-02 | -0.008 | 0.031 | 8.04E-01 | 4.02E-01 | -0.0365 | 0.0096 | -3.79 | 1.51E-04 | ---- | 6.23 | 1.01E-01 | 4 | 4,801 |
| betacryptoxanthin | 0.079 | 0.040 | 0.117 | 6.92E-05 | 8.59E-03 | 0.020 | 0.029 | 0.015 | 5.35E-02 | 2.68E-02 | NA | NA | NA | NA | NA | NA | NA | NA | 0.0472 | 0.0120 | 3.94 | 8.11E-05 | ++?? | 4.04 | 4.45E-02 | 2 | 2,958 |
| gammaCEHCglucuronide | -0.076 | -0.115 | -0.036 | 1.71E-04 | 1.18E-02 | 0.020 | -0.029 | 0.013 | 2.40E-02 | 1.20E-02 | -0.019 | 0.015 | 2.21E-01 | 1.10E-01 | -0.002 | 0.028 | 9.52E-01 | 4.76E-01 | -0.0317 | 0.0085 | -3.73 | 1.88E-04 | ---- | 6.67 | 8.32E-02 | 4 | 4,801 |
| glucose | -0.159 | -0.227 | -0.091 | 6.06E-06 | 1.88E-03 | 0.035 | -0.007 | 0.013 | 6.05E-01 | 3.03E-01 | -0.030 | 0.016 | 5.77E-02 | 2.89E-02 | -0.073 | 0.029 | 1.16E-02 | 5.80E-03 | -0.0317 | 0.0091 | -3.47 | 5.25E-04 | ---- | 18.79 | 3.01E-04 | 4 | 4,801 |
| mannitolsorbitol | -0.071 | -0.111 | -0.031 | 5.59E-04 | 3.16E-02 | 0.021 | -0.011 | 0.013 | 4.16E-01 | 2.08E-01 | -0.024 | 0.015 | 1.16E-01 | 5.81E-02 | -0.037 | 0.028 | 1.88E-01 | 9.40E-02 | -0.0278 | 0.0085 | -3.25 | 1.14E-03 | ---- | 6.25 | 1.00E-01 | 4 | 4,801 |
| mannose | -0.140 | -0.203 | -0.078 | 1.17E-05 | 2.41E-03 | 0.032 | -0.019 | 0.013 | 1.49E-01 | 7.45E-02 | -0.038 | 0.016 | 1.96E-02 | 9.80E-03 | -0.039 | 0.029 | 1.85E-01 | 9.25E-02 | -0.0369 | 0.0092 | -4.03 | 5.65E-05 | ---- | 12.32 | 6.36E-03 | 4 | 4,801 |
| Nacetylleucine | -0.112 | -0.169 | -0.056 | 1.11E-04 | 9.21E-03 | 0.029 | 0.008 | 0.013 | 5.73E-01 | 7.14E-01 | 0.006 | 0.016 | 7.02E-01 | 6.49E-01 | 0.005 | 0.029 | 8.53E-01 | 5.74E-01 | -0.0046 | 0.0091 | -0.51 | 6.13E-01 | -+++ | 15.25 | 1.62E-03 | 4 | 4,801 |
| ribitol | -0.108 | -0.171 | -0.046 | 7.12E-04 | 3.68E-02 | 0.032 | -0.019 | 0.013 | 1.53E-01 | 7.65E-02 | -0.037 | 0.016 | 2.07E-02 | 1.04E-02 | -0.047 | 0.029 | 1.01E-01 | 5.03E-02 | -0.0350 | 0.0091 | -3.84 | 1.24E-04 | ---- | 6.95 | 7.34E-02 | 4 | 4,801 |
| tyramineOsulfate | -0.066 | -0.099 | -0.033 | 1.11E-04 | 9.21E-03 | 0.017 | -0.013 | 0.013 | 3.26E-01 | 1.63E-01 | NA | NA | NA | NA | NA | NA | NA | NA | -0.0324 | 0.0103 | -3.14 | 1.71E-03 | --?? | 6.10 | 1.35E-02 | 2 | 2,958 |
| N-acetylisoleucine | -0.125 | -0.178 | -0.072 | 3.78E-06 | 1.88E-03 | 0.027 | NA | NA | NA | NA | 0.002 | 0.016 | 8.96E-01 | 5.52E-01 | -0.015 | 0.029 | 6.11E-01 | 3.05E-01 | -0.0280 | 0.0125 | -2.25 | 2.44E-02 | -?+- | 16.59 | 2.50E-04 | 3 | 2,579 |

**Abbreviations:** *BPRHS* , Boston Puerto-Rican Health Study; *HCHS/SOL* , Hispanic Community Health Study/ Study of Latinos; *ARIC* , The Atherosclerosis Risk in Communities Study; CI, Confidence Interval; SE, standard error.

Supplementary Table 6: Two-sample Mendelian randomization results for metabolites (exposure), measured in either U.S. Hispanics/Latinos from HCHS/SOL or in Europeans from Long et al. 2017, and general cognitive function (outcome) measured in Europeans from Davies et al. 2018.

| | | | | | | | Heterogeneity | | | Horizontal pleiotropy | | | |
|---|---|---|---|---|---|---|---|---|---|---|---|---|---|
| | **Exposure (metabolite)** | **Method** | **number of snps** | **Beta** | **SE** | **P-value** | **Q** | **Q_df** | **Q_pval** | **egger_intercept** | **se** | **pval** | **$R^2$ (%)** |
| Metabolites GWAS summary statistics from HCHS/SOL | betacryptoxanthin | Wald ratio | 1 | -5.28E-06 | 6.35E-06 | 4.05E-01 | | | | | | | 0.94 |
| | gammaCEHCglucuronide | Wald ratio | 1 | -3.37E-06 | 9.69E-06 | 7.28E-01 | | | | | | | 0.68 |
| | mannose | IVW | 5 | -2.81E-06 | 3.29E-06 | 3.93E-01 | 14.01 | 4 | 0.007 | | | | 12.48 |
| | mannose | MR Egger | 5 | 8.17E-07 | 5.48E-06 | 8.91E-01 | 11.31 | 3 | 0.010 | -2.05E-03 | 2.43E-03 | 4.60E-01 | |
| | mannose | IVW multiplicative random effects model | 8 | -2.18E-06 | 2.48E-06 | 3.80E-01 | 16.03 | 7 | 0.025 | | | | |
| | mannose | Weighted median | 5 | -1.72E-06 | 1.87E-06 | 3.59E-01 | | | | | | | |
| | methylthioadenosineMTA | IVW multiplicative random effects model | 2 | -5.79E-06 | 6.52E-06 | 3.74E-01 | 1.68 | 1 | 0.195 | | | | 1.97 |
| | ribitol | IVW multiplicative random effects model | 2 | 3.72E-06 | 9.67E-06 | 7.00E-01 | 2.75 | 1 | 0.097 | | | | 3.69 |
| Metabolites GWAS summary statistics from Long et al. 2017 | 5-methylthioadenosine(MTA) | Wald ratio | 1 | -2.10E-01 | 1.22E-01 | 8.36E-02 | | | | | | | |
| | glucose | Wald ratio | 1 | 2.48E-01 | 2.55E-01 | 3.32E-01 | | | | | | | |
| | mannose | Wald ratio | 1 | 3.35E-02 | 6.45E-02 | 6.03E-01 | | | | | | | |
| | ribitol | IVW | 2 | 8.63E-02 | 7.07E-02 | 2.23E-01 | 1.19 | 1 | 0.275 | | | | |
| | ribitol | IVW multiplicative random effects model | 3 | 7.14E-02 | 5.10E-02 | 1.62E-01 | 1.50 | 2 | 0.473 | | | | |

**Abbreviations:** *IVW,* Inverse variance weighted; *HCHS/SOL* , Hispanic Community Health Study/ Study of Latinos; $R^2$ , a parameter of the variance in the exposure explained by the genetic variants.

Supplementary Table 7: Two-sample Mendelian randomization results for metabolites (exposure), measured in either U.S. Hispanics/Latinos from HCHS/SOL or in Europeans from Lee et al. 2018, and general cognitive performance (outcome) measured in Europeans.

| | | | | | | | Heterogeneity | | | Horizontal pleiotropy | | | |
|---|---|---|---|---|---|---|---|---|---|---|---|---|---|
| | **Exposure (metabolite)** | **Method** | **number of snps** | **Beta** | **SE** | **P-value** | **Q** | **Q_df** | **Q_pval** | **egger_intercept** | **se** | **pval** | **$R^2$ (%)** |
| Metabolites GWAS summary statistics from HCHS/SOL | betacryptoxanthin | Wald ratio | 1 | -2.08E-05 | 1.43E-05 | 1.46E-01 | | | | | | | 0.94 |
| | gammaCEHCglucuronide | IVW multiplicative random effects model | 2 | 1.17E-05 | 7.94E-06 | 1.42E-01 | 1.10 | 1 | 2.943E-01 | | | | 3.13 |
| | mannose | Inverse variance weighted | 6 | 5.25E-06 | 6.33E-06 | 4.07E-01 | 21.73 | 5 | 5.892E-04 | | | | 12.43 |
| | mannose | MR Egger | 6 | 5.06E-06 | 1.22E-05 | 7.01E-01 | 21.73 | 4 | 2.270E-04 | 1.24E-04 | 6.48E-03 | 9.86E-01 | |
| | mannose | IVW multiplicative random effects model | 8 | 4.41E-06 | 5.54E-06 | 4.26E-01 | 26.68 | 7 | 3.810E-04 | | | | |
| | mannose | Weighted median | 6 | 4.90E-06 | 3.34E-06 | 1.42E-01 | | | | | | | |
| | methylthioadenosineMTA | Random | 2 | 2.05E-06 | 9.16E-06 | 8.23E-01 | 1.08 | 1 | 2.987E-01 | | | | 1.97 |
| | ribitol | IVW multiplicative random effects model | 2 | -2.34E-05 | 1.09E-05 | 3.22E-02 | 0.90 | 1 | 3.428E-01 | | | | 3.69 |
| Metabolites GWAS summary statistics from Long et al. 2017 | 5-methylthioadenosine(MTA) | Wald ratio | 1 | 1.52E-01 | 2.08E-01 | 4.65E-01 | | | | | | | |
| | glucose | Wald ratio | 1 | 1.85E-01 | 4.64E-01 | 6.89E-01 | | | | | | | |
| | mannose | Wald ratio | 1 | -1.27E-01 | 1.08E-01 | 2.37E-01 | | | | | | | |
| | ribitol | Inverse variance weighted | 3 | -2.42E-03 | 1.57E-01 | 9.88E-01 | 0.73 | 2 | 6.93E-01 | | | | |
| | ribitol | MR Egger | 3 | 1.58E-01 | 5.89E-01 | 8.33E-01 | 0.65 | 1 | 4.19E-01 | -2.36E-03 | 8.36E-03 | 8.25E-01 | |
| | ribitol | IVW multiplicative random effects model | 3 | -2.42E-03 | 9.50E-02 | 9.80E-01 | 0.73 | 2 | 6.93E-01 | | | | |
| | ribitol | Weighted median | 3 | 1.69E-02 | 1.86E-01 | 9.28E-01 | | | | | | | |

**Abbreviations:** *IVW,* Inverse variance weighted; *HCHS/SOL* , Hispanic Community Health Study/ Study of Latinos; $R^2$ , a parameter of the variance in the exposure explained by the genetic variants.

Supplementary Table 8: Mediterranean diet and diet scores associations with replicated/generalized Metabolites in the total HCHS/SOL analytic sample. Models adjusted for age, sex, study center, education, BMI, smoking, APOE-ε4 carrier status, smoking, and self-identified Hispanic/Latino background.

| | | Metabolites | | | | | | | |
|---|---|---|---|---|---|---|---|---|---|
| **Dietary score** | | **methylthioadenosine MTA** | **betacryptoxanthin** | **gammaCEHCglucuronide** | **glucose** | **mannitolsorbitol** | **mannose** | **ribitol** | **tyramineOsulfate** |
| **Mediterranean diet score** * | **Beta** | -0.025 | 0.122 | -0.020 | -0.030 | -0.008 | -0.050 | -0.023 | -0.054 |
| | **SE** | 0.013 | 0.012 | 0.014 | 0.014 | 0.013 | 0.013 | 0.013 | 0.014 |
| | **P-val** | 4.58E-02 | 2.80E-25 | 1.55E-01 | 2.84E-02 | 5.70E-01 | 1.89E-04 | 8.08E-02 | 6.29E-05 |
| **Legume** † | **Beta** | -0.052 | 0.033 | 0.106 | 0.072 | -0.038 | 0.022 | 0.027 | -0.037 |
| | **SE** | 0.039 | 0.036 | 0.042 | 0.042 | 0.041 | 0.041 | 0.041 | 0.042 |
| | **P-val** | 1.87E-01 | 3.51E-01 | 1.21E-02 | 8.86E-02 | 3.49E-01 | 5.85E-01 | 5.11E-01 | 3.76E-01 |
| **Fruit** † | **Beta** | -0.040 | 0.380 | -0.144 | -0.103 | -0.058 | -0.128 | -0.011 | -0.064 |
| | **SE** | 0.039 | 0.036 | 0.042 | 0.042 | 0.041 | 0.041 | 0.041 | 0.042 |
| | **P-val** | 3.05E-01 | 2.69E-26 | 6.75E-04 | 1.50E-02 | 1.61E-01 | 1.92E-03 | 7.92E-01 | 1.30E-01 |
| **Vegtables** † | **Beta** | -0.046 | 0.208 | -0.032 | 0.017 | 0.039 | -0.020 | -0.014 | -0.022 |
| | **SE** | 0.040 | 0.037 | 0.043 | 0.043 | 0.042 | 0.042 | 0.042 | 0.043 |
| | **P-val** | 2.54E-01 | 1.20E-08 | 4.58E-01 | 6.89E-01 | 3.48E-01 | 6.35E-01 | 7.33E-01 | 6.13E-01 |
| **Whole grain** † | **Beta** | -0.059 | 0.137 | -0.064 | 0.072 | 0.083 | 0.109 | -0.025 | -0.099 |
| | **SE** | 0.044 | 0.040 | 0.048 | 0.048 | 0.046 | 0.046 | 0.047 | 0.047 |
| | **P-val** | 1.87E-01 | 6.78E-04 | 1.79E-01 | 1.33E-01 | 7.35E-02 | 1.93E-02 | 5.91E-01 | 3.71E-02 |
| **Nuts** † | **Beta** | -0.085 | 0.085 | -0.050 | -0.133 | -0.111 | -0.135 | -0.161 | -0.071 |
| | **SE** | 0.055 | 0.050 | 0.060 | 0.059 | 0.058 | 0.058 | 0.058 | 0.059 |
| | **P-val** | 1.23E-01 | 9.24E-02 | 4.05E-01 | 2.53E-02 | 5.38E-02 | 1.97E-02 | 5.69E-03 | 2.29E-01 |
| **Red meat** † | **Beta** | 0.074 | 0.094 | 0.034 | -0.144 | 0.050 | -0.142 | 0.026 | -0.071 |
| | **SE** | 0.040 | 0.036 | 0.043 | 0.043 | 0.042 | 0.042 | 0.042 | 0.043 |
| | **P-val** | 6.57E-02 | 9.53E-03 | 4.25E-01 | 8.28E-04 | 2.30E-01 | 7.15E-04 | 5.29E-01 | 9.44E-02 |
| **Fish** † | **Beta** | -0.045 | 0.123 | 0.001 | 0.001 | -0.009 | 0.017 | -0.041 | -0.065 |
| | **SE** | 0.042 | 0.039 | 0.046 | 0.046 | 0.044 | 0.044 | 0.045 | 0.045 |
| | **P-val** | 2.86E-01 | 1.52E-03 | 9.87E-01 | 9.83E-01 | 8.47E-01 | 7.02E-01 | 3.52E-01 | 1.52E-01 |
| **Mufa sfa** † | **Beta** | 0.026 | -0.045 | 0.038 | -0.043 | -0.031 | -0.067 | -0.019 | -0.014 |
| | **SE** | 0.039 | 0.036 | 0.042 | 0.042 | 0.041 | 0.041 | 0.041 | 0.042 |
| | **P-val** | 5.04E-01 | 2.02E-01 | 3.69E-01 | 3.05E-01 | 4.53E-01 | 1.02E-01 | 6.45E-01 | 7.38E-01 |
| **Alcohol** † | **Beta** | 0.125 | 0.013 | -0.169 | -0.145 | 0.105 | -0.327 | 0.035 | -0.049 |
| | **SE** | 0.087 | 0.079 | 0.093 | 0.093 | 0.091 | 0.091 | 0.091 | 0.092 |
| | **P-val** | 1.49E-01 | 8.74E-01 | 7.03E-02 | 1.19E-01 | 2.48E-01 | 3.18E-04 | 7.01E-01 | 5.97E-01 |

* Summary results from model 1 for the Mediterranean diet score.

† Summary results from model 2 for each of the 9 dietary scores.

Supplementary Table 9: Two-sample Mendelian randomization results for association between single-food intake scores measured in Europeans from UK Biobank and general cognitive function measured in Europeans from Lee et al. 2017.

| dietary scores (UK Biobank) and cognitive performance | | | | | | Heterogeneity | | | Horizontal pleiotropy | | |
|---|---|---|---|---|---|---|---|---|---|---|---|
| **exposure** | **method** | **nsnp** | **Beta** | **SE** | **P-value** | **Q** | **Q_df** | **Q_pval** | **egger_intercept** | **se** | **pval** |
| alcohol | IVW | 74 | -2.06E-01 | 5.74E-02 | 3.36E-04 | 586.9 | 73 | 2.59E-81 | | | |
| | IVW multiplicative random effects model | 75 | -1.99E-01 | 5.75E-02 | 5.34E-04 | 601.9 | 74 | 1.03E-83 | | | |
| | MR Egger | 74 | 5.48E-02 | 1.08E-01 | 6.14E-01 | 529.7 | 72 | 6.71E-71 | -5.59E-03 | 2.00E-03 | 6.77E-03 |
| | Weighted median | 74 | 3.27E-02 | 4.11E-02 | 4.26E-01 | | | | | | |
| beef | IVW | 15 | 1.92E-01 | 1.69E-01 | 2.57E-01 | 132.2 | 14 | 2.50E-21 | | | |
| | IVW multiplicative random effects model | 15 | 1.92E-01 | 1.69E-01 | 2.57E-01 | 132.2 | 14 | 2.50E-21 | | | |
| | MR Egger | 15 | -2.15E-02 | 7.01E-01 | 9.76E-01 | 131.2 | 13 | 1.21E-21 | 3.43E-03 | 1.09E-02 | 7.58E-01 |
| | Weighted median | 15 | 2.02E-01 | 1.20E-01 | 9.16E-02 | | | | | | |
| cookedveg | IVW | 23 | 6.65E-02 | 9.04E-02 | 4.62E-01 | 86.5 | 22 | 1.33E-09 | | | |
| | IVW multiplicative random effects model | 25 | 8.65E-02 | 8.68E-02 | 3.19E-01 | 93.0 | 24 | 4.57E-10 | | | |
| | MR Egger | 23 | -1.21E-01 | 4.44E-01 | 7.89E-01 | 85.8 | 21 | 8.63E-10 | 2.77E-03 | 6.44E-03 | 6.71E-01 |
| | Weighted median | 23 | 1.13E-01 | 7.48E-02 | 1.31E-01 | | | | | | |
| driedfruit | IVW | 31 | -3.16E-01 | 1.30E-01 | 1.49E-02 | 358.7 | 30 | 5.77E-58 | | | |
| | IVW multiplicative random effects model | 33 | -3.14E-01 | 1.23E-01 | 1.06E-02 | 358.8 | 32 | 6.43E-57 | | | |
| | MR Egger | 31 | 9.41E-01 | 4.25E-01 | 3.48E-02 | 270.5 | 29 | 5.16E-41 | -2.05E-02 | 6.67E-03 | 4.56E-03 |
| | Weighted median | 31 | -2.18E-01 | 7.40E-02 | 3.24E-03 | | | | | | |
| freshfruit | IVW | 77 | -8.33E-02 | 5.94E-02 | 1.61E-01 | 532.8 | 76 | 9.63E-70 | | | |
| | IVW multiplicative random effects model | 82 | -9.17E-02 | 5.80E-02 | 1.14E-01 | 572.1 | 81 | 5.67E-75 | | | |
| | MR Egger | 77 | -3.45E-02 | 2.05E-01 | 8.67E-01 | 532.3 | 75 | 4.35E-70 | -8.11E-04 | 3.26E-03 | 8.04E-01 |
| | Weighted median | 77 | 1.59E-02 | 4.14E-02 | 7.02E-01 | | | | | | |
| lambmutton | IVW | 27 | -9.77E-02 | 1.35E-01 | 4.71E-01 | 267.1 | 26 | 7.42E-42 | | | |
| | IVW multiplicative random effects model | 27 | -9.77E-02 | 1.35E-01 | 4.71E-01 | 267.1 | 26 | 7.42E-42 | | | |
| | MR Egger | 27 | 9.68E-01 | 7.49E-01 | 2.08E-01 | 246.5 | 25 | 2.66E-38 | -1.63E-02 | 1.13E-02 | 1.61E-01 |
| | Weighted median | 27 | 9.39E-02 | 7.84E-02 | 2.31E-01 | | | | | | |
| nonoilyfish | IVW | 11 | 1.19E-01 | 9.74E-02 | 2.22E-01 | 24.1 | 10 | 7.35E-03 | | | |
| | IVW multiplicative random effects model | 11 | 1.19E-01 | 9.74E-02 | 2.22E-01 | 24.1 | 10 | 7.35E-03 | | | |
| | MR Egger | 11 | -7.34E-01 | 4.04E-01 | 1.03E-01 | 15.9 | 9 | 6.96E-02 | 1.31E-02 | 6.08E-03 | 5.92E-02 |
| | Weighted median | 11 | -1.91E-02 | 9.46E-02 | 8.40E-01 | | | | | | |
| oilyfish | IVW | 56 | 8.64E-02 | 6.39E-02 | 1.77E-01 | 313.1 | 55 | 8.47E-38 | | | |
| | IVW multiplicative random effects model | 58 | 1.19E-01 | 6.82E-02 | 7.99E-02 | 379.2 | 57 | 4.12E-49 | | | |
| | MR Egger | 56 | 5.46E-01 | 2.67E-01 | 4.59E-02 | 295.9 | 54 | 4.34E-35 | -7.43E-03 | 4.20E-03 | 8.24E-02 |
| | Weighted median | 56 | 6.56E-03 | 5.00E-02 | 8.96E-01 | | | | | | |
| pork | IVW | 13 | -2.41E-01 | 1.47E-01 | 1.01E-01 | 64.6 | 12 | 3.20E-09 | | | |
| | IVW multiplicative random effects model | 14 | -2.81E-01 | 1.39E-01 | 4.24E-02 | 68.9 | 13 | 1.28E-09 | | | |
| | MR Egger | 13 | -7.69E-02 | 6.13E-01 | 9.03E-01 | 64.2 | 11 | 1.54E-09 | -2.45E-03 | 8.86E-03 | 7.88E-01 |
| | Weighted median | 13 | -1.50E-01 | 9.94E-02 | 1.32E-01 | | | | | | |
| poultry | IVW | 4 | 2.81E-02 | 2.48E-01 | 9.10E-01 | 14.6 | 3 | 2.19E-03 | | | |
| | IVW multiplicative random effects model | 4 | 2.81E-02 | 2.48E-01 | 9.10E-01 | 14.6 | 3 | 2.19E-03 | | | |
| | MR Egger | 4 | -4.60E+00 | 2.29E+00 | 1.82E-01 | 4.8 | 2 | 9.20E-02 | 6.20E-02 | 3.06E-02 | 1.80E-01 |
| | Weighted median | 4 | -1.51E-01 | 1.48E-01 | 3.07E-01 | | | | | | |
| processmeat | IVW | 19 | -8.43E-02 | 1.83E-01 | 6.44E-01 | 223.6 | 18 | 1.86E-37 | | | |
| | IVW multiplicative random effects model | 19 | -8.43E-02 | 1.83E-01 | 6.44E-01 | 223.6 | 18 | 1.86E-37 | | | |
| | MR Egger | 19 | -9.49E-01 | 9.68E-01 | 3.40E-01 | 213.2 | 17 | 6.38E-36 | 1.28E-02 | 1.40E-02 | 3.75E-01 |
| | Weighted median | 19 | -7.78E-02 | 9.54E-02 | 4.15E-01 | | | | | | |
| rawveg | IVW | 32 | 8.75E-02 | 6.98E-02 | 2.10E-01 | 110.4 | 31 | 7.79E-11 | | | |
| | IVW multiplicative random effects model | 33 | 9.01E-02 | 6.78E-02 | 1.84E-01 | 110.6 | 32 | 1.41E-10 | | | |
| | MR Egger | 32 | 9.09E-02 | 3.09E-01 | 7.71E-01 | 110.4 | 30 | 3.98E-11 | -5.52E-05 | 4.91E-03 | 9.91E-01 |
| | Weighted median | 32 | 1.67E-01 | 6.04E-02 | 5.71E-03 | | | | | | |

**Abbreviations:** *IVW,* Inverse variance weighted.

Supplementary Table 10: Two-sample Mendelian randomization results for association between general cognitive function measured in Europeans from Lee et al. 2017 and single-food intake scores measured in Europeans from UK Biobank.

| Cognitive performance (Lee JJ, GWAS catalog) and dietary scores (UK Biobank) | | | | | | Heterogeneity | | | Horizontal pleiotropy | | |
|---|---|---|---|---|---|---|---|---|---|---|---|
| Outcome | method | nsnp | Beta | SE | P-value | Q | Q_df | Q_pval | egger_intercept | se | pval |
| alcohol | IVW | 70 | -1.33E-01 | 2.62E-02 | 3.60E-07 | 263.7 | 69 | 1.53E-24 | | | |
| | IVW multiplicative random effects model | 162 | -1.61E-01 | 1.90E-02 | 2.06E-17 | 760.7 | 161 | 1.36E-78 | | | |
| | MR Egger | 70 | -2.46E-01 | 1.07E-01 | 2.42E-02 | 259.2 | 68 | 4.19E-24 | 2.46E-03 | 2.26E-03 | 2.81E-01 |
| | Weighted median | 70 | -8.91E-02 | 2.44E-02 | 2.58E-04 | | | | | | |
| beef | IVW | 70 | 5.44E-02 | 1.95E-02 | 5.18E-03 | 143.0 | 69 | 4.23E-07 | | | |
| | IVW multiplicative random effects model | 162 | 5.24E-02 | 1.33E-02 | 8.14E-05 | 365.8 | 161 | 5.64E-18 | | | |
| | MR Egger | 70 | 2.94E-02 | 7.97E-02 | 7.14E-01 | 142.8 | 68 | 3.05E-07 | 5.47E-04 | 1.69E-03 | 7.47E-01 |
| | Weighted median | 70 | 3.55E-02 | 2.09E-02 | 9.01E-02 | | | | | | |
| cookedveg | IVW | 70 | 7.22E-02 | 2.18E-02 | 9.10E-04 | 177.8 | 69 | 1.52E-11 | | | |
| | IVW multiplicative random effects model | 162 | 4.86E-02 | 1.48E-02 | 1.05E-03 | 452.1 | 161 | 1.87E-29 | | | |
| | MR Egger | 70 | 1.07E-01 | 8.91E-02 | 2.35E-01 | 177.3 | 68 | 1.06E-11 | -7.58E-04 | 1.89E-03 | 6.90E-01 |
| | Weighted median | 70 | 6.49E-02 | 2.28E-02 | 4.37E-03 | | | | | | |
| driedfruit | IVW | 70 | -7.46E-02 | 2.14E-02 | 5.02E-04 | 173.0 | 69 | 6.68E-11 | | | |
| | IVW multiplicative random effects model | 162 | -8.37E-02 | 1.58E-02 | 1.16E-07 | 514.1 | 161 | 1.66E-38 | | | |
| | MR Egger | 70 | 3.34E-02 | 8.69E-02 | 7.02E-01 | 168.9 | 68 | 1.47E-10 | -2.36E-03 | 1.84E-03 | 2.04E-01 |
| | Weighted median | 70 | -8.30E-02 | 2.23E-02 | 2.03E-04 | | | | | | |
| freshfruit | IVW | 70 | -3.65E-02 | 2.25E-02 | 1.05E-01 | 193.4 | 69 | 9.82E-14 | 9.66E-04 | 1.96E-03 | 6.23E-01 |
| | IVW multiplicative random effects model | 162 | -2.81E-02 | 1.79E-02 | 1.17E-01 | 673.5 | 161 | 7.83E-64 | | | |
| | MR Egger | 70 | -8.06E-02 | 9.21E-02 | 3.84E-01 | 192.7 | 68 | 7.24E-14 | | | |
| | Weighted median | 70 | -2.94E-02 | 2.14E-02 | 1.70E-01 | | | | | | |
| lambmutton | IVW | 70 | -3.39E-02 | 2.13E-02 | 1.11E-01 | 169.9 | 69 | 1.78E-10 | | | |
| | IVW multiplicative random effects model | 162 | -3.66E-02 | 1.41E-02 | 9.31E-03 | 408.3 | 161 | 1.92E-23 | | | |
| | MR Egger | 70 | -1.07E-01 | 8.67E-02 | 2.22E-01 | 168.0 | 68 | 1.97E-10 | 1.60E-03 | 1.84E-03 | 3.88E-01 |
| | Weighted median | 70 | -6.95E-02 | 2.17E-02 | 1.37E-03 | | | | | | |
| nonoilyfish | IVW | 70 | 5.29E-02 | 1.71E-02 | 1.98E-03 | 109.6 | 69 | 1.35E-03 | | | |
| | IVW multiplicative random effects model | 162 | 4.60E-02 | 1.16E-02 | 7.33E-05 | 276.5 | 161 | 4.01E-08 | | | |
| | MR Egger | 70 | 5.40E-03 | 6.99E-02 | 9.39E-01 | 108.8 | 68 | 1.23E-03 | 1.04E-03 | 1.48E-03 | 4.85E-01 |
| | Weighted median | 70 | 6.86E-02 | 2.02E-02 | 6.74E-04 | | | | | | |
| oilyfish | IVW | 70 | 2.05E-03 | 2.45E-02 | 9.33E-01 | 225.6 | 69 | 1.61E-18 | | | |
| | IVW multiplicative random effects model | 162 | -2.87E-02 | 1.61E-02 | 7.56E-02 | 537.9 | 161 | 4.05E-42 | | | |
| | MR Egger | 70 | 5.62E-03 | 1.00E-01 | 9.56E-01 | 225.6 | 68 | 8.77E-19 | -7.81E-05 | 2.13E-03 | 9.71E-01 |
| | Weighted median | 70 | -1.62E-02 | 2.29E-02 | 4.79E-01 | | | | | | |
| pork | IVW | 70 | 2.17E-02 | 1.99E-02 | 2.75E-01 | 147.9 | 69 | 1.10E-07 | | | |
| | IVW multiplicative random effects model | 162 | 2.11E-02 | 1.23E-02 | 8.65E-02 | 311.5 | 161 | 1.14E-11 | | | |
| | MR Egger | 70 | -4.79E-02 | 8.10E-02 | 5.56E-01 | 146.2 | 68 | 1.18E-07 | 1.52E-03 | 1.72E-03 | 3.79E-01 |
| | Weighted median | 70 | 1.74E-02 | 2.23E-02 | 4.35E-01 | | | | | | |
| poultry | IVW | 70 | 6.62E-02 | 1.96E-02 | 7.49E-04 | 145.0 | 69 | 2.46E-07 | | | |
| | IVW multiplicative random effects model | 162 | 5.28E-02 | 1.29E-02 | 4.32E-05 | 343.0 | 161 | 3.24E-15 | | | |
| | MR Egger | 70 | 1.28E-01 | 8.02E-02 | 1.15E-01 | 143.7 | 68 | 2.39E-07 | -1.35E-03 | 1.70E-03 | 4.30E-01 |
| | Weighted median | 70 | 6.48E-02 | 2.21E-02 | 3.41E-03 | | | | | | |
| processmeat | IVW | 70 | -1.93E-02 | 2.10E-02 | 3.58E-01 | 167.1 | 69 | 4.08E-10 | | | |
| | IVW multiplicative random effects model | 162 | -1.09E-02 | 1.38E-02 | 4.27E-01 | 391.6 | 161 | 3.07E-21 | | | |
| | MR Egger | 70 | -2.90E-02 | 8.63E-02 | 7.38E-01 | 167.1 | 68 | 2.59E-10 | 2.12E-04 | 1.83E-03 | 9.08E-01 |
| | Weighted median | 70 | -8.30E-03 | 2.18E-02 | 7.04E-01 | | | | | | |
| rawveg | IVW | 70 | 7.47E-02 | 2.30E-02 | 1.14E-03 | 198.0 | 69 | 2.18E-14 | | | |
| | IVW multiplicative random effects model | 162 | 5.38E-02 | 1.47E-02 | 2.47E-04 | 442.7 | 161 | 3.87E-28 | | | |
| | MR Egger | 70 | 1.27E-01 | 9.39E-02 | 1.82E-01 | 197.0 | 68 | 1.73E-14 | -1.14E-03 | 1.99E-03 | 5.70E-01 |
| | Weighted median | 70 | 3.09E-02 | 2.24E-02 | 1.68E-01 | | | | | | |

**Abbreviations:** *IVW,* Inverse variance weighted.
